# African ancestry neurodegeneration risk variant disrupts an intronic branchpoint in GBA1

**DOI:** 10.1101/2024.02.20.24302827

**Authors:** Pilar Álvarez Jerez, Peter A. Wild Crea, Daniel M. Ramos, Emil K. Gustavsson, Mandy Radefeldt, Mary B. Makarious, Oluwadamilola O. Ojo, Kimberley J. Billingsley, Laksh Malik, Kensuke Daida, Sarah Bromberek, Carol Hu, Zachary Schneider, Aditya L. Surapaneni, Julia Stadler, Mie Rizig, Huw R. Morris, Caroline B. Pantazis, Hampton L. Leonard, Laurel Screven, Yue A. Qi, Mike A. Nalls, Sara Bandres-Ciga, John Hardy, Henry Houlden, Celeste Eng, Esteban González Burchard, Linda Kachuri, Global Parkinson’s Genetics Program (GP2), Andrew B. Singleton, Steffen Fischer, Peter Bauer, Xylena Reed, Mina Ryten, Christian Beetz, Michael Ward, Njideka U. Okubadejo, Cornelis Blauwendraat

## Abstract

Recently, a novel African ancestry specific Parkinson’s disease (PD) risk signal was identified at the gene encoding glucocerebrosidase (*GBA1*). This variant (rs3115534-G) is carried by ∼50% of West African PD cases and imparts a dose-dependent increase in risk for disease. The risk variant has varied frequencies across African ancestry groups, but is almost absent in European and Asian ancestry populations. *GBA1* is a gene of high clinical and therapeutic interest. Damaging bi-allelic protein-coding variants cause Gaucher disease and mono-allelic variants confer risk for PD and Dementia with Lewy Bodies, likely by reducing the function of glucocerebrosidase. Interestingly, the novel African ancestry specific *GBA1* risk variant is a non-coding variant, suggesting a different mechanism of action. Using full length RNA transcript sequencing, we identified intron 8 expression in risk variant carriers (G) but not in non-variant carriers (T). Antibodies targeting the N-terminus of glucocerebrosidase showed that this intron-retained isoform is likely not protein coding and subsequent proteomics did not identify a shorter protein isoform, suggesting the disease mechanism is RNA-based. CRISPR editing of the reported index variant (rs3115534) revealed that this is the responsible sequence alteration driving production of these intron 8 containing transcripts. Follow-up analysis of this variant showed that it is in a key intronic branchpoint sequence and therefore has important implications in splicing and disease. In addition, when measuring glucocerebrosidase activity we identified a dose-dependent reduction in risk variant carriers (G). Overall, we report the functional effect of a *GBA1* non-coding risk variant, which acts by interfering with the splicing of functional *GBA1* transcripts, resulting in reduced protein levels and reduced glucocerebrosidase activity. This understanding reveals a novel therapeutic target in an underserved and underrepresented population.

## Introduction

Dementia with Lewy Bodies (DLB) and Parkinson’s disease (PD) are believed to be caused by a combination of aging, environmental factors, and genetics. Genetics has provided valuable insights into the underlying biology of disease. Damaging variants in multiple genes have been shown to cause disease and numerous variants have been associated with increased risk ^1–3^. One particular gene of interest is *GBA1* (previously known as *GBA*) which encodes the lysosomal enzyme glucocerebrosidase (GCase). Damaging coding variants in *GBA1* increase the risk for PD and DLB across a wide odds ratio spectrum ^4,5^. Interestingly, the phenotype of individuals with PD carrying *GBA1* variants is characterized by faster progression and a higher frequency of dementia compared to non-carriers ^6,7^. It is most commonly hypothesized that *GBA1* mutations confer risk by reducing GCase activity. Furthermore, damaging *GBA1* variants are enriched in certain populations e.g., p.E326K in Northern Europeans and p.N370S in Ashkenazi Jews ^8^. Bi-allelic *GBA1* variants cause Gaucher disease, a lysosomal storage disease that leads to a variety of clinical presentations.

In the human genome, *GBA1* is located on chromosome 1q22 and is adjacent to its pseudogene, known as *GBAP1*. *GBAP1* has very high sequence homology with *GBA1* (96%). Consequently, accurately mapping short read RNA and DNA sequencing in this region is a complex task ^9,10^. Much is unknown about the potential function of *GBAP1*, but using long-read sequencing, which overcomes mapping issues in this genomic region, it is clear that *GBAP1* is expressed at the RNA level and there is some evidence for protein expression ^10^.

Recently, a novel PD *GBA1* risk signal was identified in the first African ancestry PD GWAS ^11^. The main index variant was remarkably common in West African populations with an estimated frequency of ∼50% in West African PD cases and a reported odds ratio (OR) of 1.58 (95% CI = 1.37 – 1.80, P=2.397E-14) per allele. In addition, this variant was associated with an earlier age at onset of 2 years per allele (BETA =−2.004, SE =0.57, P = 0.0005). Strikingly, the main index variant (rs3115534, NM_000157.4(GBA1):c.1225-34C>A) is a non-coding variant reported to be the strongest expression (eQTL) and protein quantitative trait locus (pQTL) for *GBA1* in the African ancestry population ^12,13^. We have shown previously that there are no common coding variants or structural variants in linkage disequilibrium with rs3115543, implying a disease mechanism independent of protein-coding or genomic structural variants at this locus ^11^.

Here, we elucidate the disease mechanism of an intronic *GBA1* PD risk variant seen as the first and major genetic risk factor in African ancestry populations. Additionally, we show that the index variant (rs3115534) causes abnormal splicing and processing of *GBA1* transcripts, only present in risk variant carriers.

## Methods

### Biosamples used for assessment of effects of GBA1 rs3115534

To assess the potential molecular downstream effects of the non-coding *GBA1* variant rs3115534, we used data from the Global Parkinson’s Genetics Program data release 5 (GP2, https://gp2.org/, DOI 10.5281/zenodo.10472143) and identified variant carriers of interest with matching Lymphoblastoid Cell Lines (LCLs) at the Coriell Institute for Medical Research (Camden, NJ, USA) (https://www.coriell.org/). In addition, we accessed brain tissue samples from the Human Brain Collection Core (HBCC) with and without the non-coding *GBA1* variant (rs3115534). For a full overview of included samples and their demographics see **Supplementary Table 1**. Note that the genome reference allele (hg38) is G for rs3115534, which is also the risk allele, and T for the alternative non-risk allele. Given that *GBA1* is transcribed from the antisense strand, C is risk and A is non-risk on the RNA level.

### DNA extraction from cell pellets and brain tissue samples

DNA was extracted from LCLs (2e6 cells) following the Pacific Biosciences (PacBio) HMW DNA Extraction Cultured Cells Protocol with the Nanobind tissue kit (PacBio, 102-203-100). The extraction occurred on a KingFisher Apex System (Thermo Fisher Scientific, 5400920). For brain tissue, 40 mg of frontal cortex tissue was cut and manually homogenized with a TissueRuptor (Qiagen, 9002755) in buffer CT (Pacbio, 102-280-300). Then, the DNA was extracted following the PacBio HMW DNA Extraction Cultured Cells Protocol with the Nanobind tissue kit (PacBio, 102-203-100) using the KingFisher Apex System (Thermo Fisher Scientific, 5400920). The DNA from LCLs and brain tissue was quantified using the Qubit dsDNA BR assay (Invitrogen, Q32850) and sized with a Femto Pulse System (Agilent, M5330AA). The samples then underwent a size selection to remove fragments under 15 kb using the Short Read Eliminator Kit (Pacbio, 102-208-300). After size selection, the DNA was sheared to a target size of 30 kb using the Megaruptor 3 (Diagenode, B060100003) with Fluid+ needles (Diagenode, E07020001) at speed 45 for 2 cycles. DNA was then quantified and resized with the Qubit dsDNA BR assay and the Femto Pulse System. Samples needed to have at least 4.5 ug of DNA and be between 20-40 kb in size to take forward into library preparation. For further details please see Cogan et al. 2023 and Billingsley et al. 2023 ^1415^.

### Oxford Nanopore Technologies DNA library preparation and sequencing

Sequencing libraries were prepared with Oxford Nanopore Technologies’ (ONT) SQK-LSK110 kit and 400 ng of prepared library per sample was loaded onto R9.4.1 flow cells on ONT’s PromethION device with Minknow 22.10.7 software. Samples were sequenced over 72 hours with 1-2 additional library loads per flow cell. Sequencing data resulted in an average coverage of 30x and an N50 of around 30 kb per sample.

### RNA extraction from cell pellets and brain tissue samples

RNA was extracted from LCLs (5e6 cells) and brain tissue (40mg) using the RNA Direct-zol miniprep kit and its corresponding protocol (Zymo Research, R2050). In short, samples were resuspended in 600uL of TRI reagent and an equal volume of 100% ethanol. Brain tissue needed additional homogenization with a Dounce homogenizer during the resuspension steps. Each mixture was then transferred into a Zymo-Spin IICR Column, centrifuged, and transferred to a new collection tube. DNase I treatment was performed using the recommended guidelines. Washing steps were performed with Zymo Research’s Direct-zol RNA PreWash and RNA Wash Buffer according to the protocol. Finally, RNA was eluted in RNase-free water and quality control was performed on Agilent’s TapeStation 4200. All cell lines had a RIN > 9 while brain RNA had RINs between 5.7 and 8.6, see **Supplementary Table 1** for additional details.

### Oxford Nanopore Technologies cDNA library preparation and sequencing

200 ng of total RNA from the LCLs and brain tissue was prepared for sequencing using ONT’s cDNA-PCR SQK-PCS111 library preparation kit and protocol with modifications. This library preparation relies on an oligo(dT)-based polyA selection. The protocol modifications included: 1) an additional bead clean up after reverse transcription with 11.25 uL of RNAse-free XP beads (Beckman Coulter, A63987), short fragment buffer washes (ONT, PCS111 Kit), and elution into 22.5 uL of elution buffer 2) PCR settings during amplification were adjusted to set the annealing steps to 12 cycles. After library preparation, total RNA was quantified using the Qubit dsDNA HS Assay Kit (Invitrogen, Q32851). Then, 22 fmol of prepared library was loaded onto R9.4.1 PromethION flow cells and sequenced for 72 hours using the Minknow 22.10.7 software.

### Single nuclei cDNA library preparation

Nuclei from frozen brain tissue were prepared using a modified homogenization protocol ^16,17^. Briefly, 30-50mg of tissue was homogenized in a Dounce homogenizer with 20 strokes of the loose pestle and 20 strokes of the tight pestle with 1x lysis buffer (10mM Tris-HCl (Sigma-Aldrich, T2194-1L), 10mM NaCl (Sigma-Aldrich, 5922C-500ML), 3mM MgCl2 (Sigma-Aldrich, M1028-100ML), 0.1% Nonidet P40 Substitute/IGEPAL CA-630 (Sigma-Aldrich, I8896-50ML), 1mM DTT (Sigma-Aldrich, 646563-10X.5ML) and 1U/ul RNase inhibitors (Sigma-Aldrich, 03335402001)). Nuclei were filtered and collected by centrifugation through a sucrose cushion gradient (Sigma-Aldrich, NUC201-1KT). Myelin and debris were removed, and nuclei washed (10mM Tris-HCl, 10mM NaCl, 3mM MgCl2, 1% Bovine Serum Albumin (Miltenyi Biotec, 130-091-376), 0.1% Tween-20 (BioRad, 1610781), 1mM DTT), pelleted and permeabilized in 0.1x lysis buffer 2 (10mM Tris-HCl, 10mM NaCl, 3mM MgCl2, 1% Bovine Serum Albumin, 0.1% Nonidet P40 Substitute/IGEPAL CA-630, 0.01% Digitonin (Invitrogen, BN20061)), washed, and counted. The resulting nuclei were then prepared using the 10x Genomics Single Cell Multiome ATAC + Gene Expression kit (10X Genomics, 1000283) and loaded on Single Cell Chip J (10x Genomics, 1000234) for recovery of 10,000 nuclei. After Tn5 transposition, single cell isolation, barcoding, and pre-amplification, cDNA was generated as directed in the user guide with polydT-primers for reverse transcription. The resulting cDNA libraries were quantified with Qubit dsDNA HS reagents (Invitrogen, Q33231) and average fragment size was determined using high sensitivity DNA D5000 screentape analysis (Agilent Technologies, 5067-5593, 5067-5592).

### GBA1 targeted transcript capture for long-read RNA sequencing

Next, to identify the cell type(s) expressing the novel intron-containing *GBA1* transcript, we performed long-read sequencing on the single cell cDNA libraries generated from the 10X Genomics Single Cell Multiome ATAC + Gene Expression kit. We first targeted *GBA1* in the single cell cDNA libraries using Pacbio’s Customer Collaboration Iso-Seq Express Capture Using IDT xGEN Lockdown Probes protocol (https://ostr.ccr.cancer.gov/wp-content/uploads/2023/06/PacBio_TargetedIso-Seq.pdf) with primer modifications to tailor it to ONT sequencing ^10^. In short, 10 ng of each single cell cDNA library was amplified with ONT cDNA 10X primers taken from ^18^ and the NEBNext Single Cell/Low Input cDNA Synthesis & Amplification Module (New England Biolabs, E6421S). After amplification, the cDNA was cleaned with ProNex beads (Promega, NG2001). A total of 500 ng of cDNA was then hybridized using custom *GBA1* xGen Lockdown Probes (IDT) (**Supplementary Table 2**) and washed with the xGen Lockdown Hybridization and Wash Kit (IDT, 1080577). The captured cDNA was then amplified using the same primers as above. For full details of reagents and PCR conditions please see **Supplementary Table 3**. After the *GBA1* capture, 10 ng of cDNA was taken into library preparation using an optimized version of the ONT PCS111 kit. The cDNA first underwent a biotin tagging reaction with custom oligos and PCR amplification, after which it was cleaned using AMPure XP beads (Beckman Coulter, A63881). Next, the cDNA was bound to M280 streptavidin beads (Invitrogen, 11205D) and amplified with the ONT cDNA Primer. The cDNA underwent AMPure XP bead cleaning once more and was quantified using the Qubit dsDNA HS reagents. 35 fmol were taken forward, the RAP T adapter was added, and the sample underwent standard loading on an R.9.4.1 flow cell on a PromethION with Minknow 22.10.7 software.

### Whole genome DNA long-read sequencing analysis

All DNA sequencing runs were basecalled on NIH’s HPC (Biowulf) using Guppy (v6.1.2) in super accuracy mode with the *dna_r9.4.1_450bps_modbases_5mc_cg_sup_prom.cfg* configuration file. Basecalled files were mapped to hg38 using Minimap2 (v2.24/2.26) ^19^ preserving methylation tags (samtools fastq -TMm,Ml ${FASTQ_PATH}/${BAM_FILE} | minimap2 -y -x map-ont -t 20 -a --eqx -k 17 -K 10g). SNVs were called using Clair3 (v1.0.4) ^20^ and structural variants called using Sniffles (v2.2) ^21^.

### Untargeted RNA long-read sequencing analysis

All RNA sequencing runs were basecalled on NIH’s HPC (Biowulf) using Guppy (v6.1.2) in super accuracy mode with the *dna_r9.4.1_450bps_sup_prom.cfg* configuration file for cDNA. Pychopper v2.7.1 ^22^ was run on the cDNA FASTQs with a minimum mean read quality of 7.0 and a minimum segment length of 50, for kit SQK-PCS111. Reads passing quality control were mapped to hg38 using Minimap2 (v2.26) ^19^ with splice aware parameters (-t 10 -ax splice -k14 - uf). Only reads with a minimum mapping quality of 40 and flagged as primary alignment were kept. For transcript calling and quantification, we used Stringtie2 (v2.2.1) ^23^. Stringtie2 was first run in long-read reference free mode (stringtie -L -R -m 50) to capture our novel intronic region. Then, we generated a reference GTF file with the canonical *GBA1* transcript (ENST00000368373.8) and the transcript containing part of intron 8, and reran our samples against this reference (stringtie --rf -G ${reference_annotation} -L -v -p 10) for quantification of the novel intron-containing transcript and canonical transcript. Each mapped bam and Stringtie2 annotation was then manually inspected on IGV (v2.16.0) ^24^.

Additionally, we calculated regional sequencing depth for different *GBA1* regions. For this, we used Samtools (v1.17) ^25^ to subset the *GBA1* regions from each hg38 mapped bam and used samtools coverage -q5 -Q20 --ff UNMAP,SECONDARY,QCFAIL,DUP -r $chr:$start-$end ${IN} to calculate the mean depth at each region of interest. These regions of interest included: 1) exons 8 and 9; 2) intron 8; 3) the ∼40 bp novel transcript region in intron 8; 4) intron 8 minus the ∼40 bp novel transcript region and 5) *GBA1* and *GBAP1*. These coordinates were based on ENST00000368373.8 for *GBA1* and the exact bed file can be found in **Supplementary Table 4**. To normalize coverage metrics across samples, we extracted the number of uniquely mapped reads per sample over the whole genome using samtools view -c -F 260 ${IN} and divided these reads by a million. We then divided our mean depth metric by the number of reads for a normalized coverage per million metric. Normalized coverage counts were averaged across rs3115534 genotypes and plotted using ggplot2 ^26^ in R (v4.3.0).

Lastly, to check that these reads map uniquely to *GBA1* and not to *GBAP1*, we manually edited the rs3115534 variant in the FASTQ data and looked for differences in mapping. To do this, we subsetted the *GBA1* and *GBAP1* regions for 1 GG and 1 TT from the hg38 aligned bam files and converted the subsets back to a FASTQ using samtools (v1.17) bam2fq. Then, using Virtual Studio Code ^27^, we manually changed the rs3115534 variant from a G to a T and vice versa, and remapped the edited FASTQ. Mapped bam files were then inspected on IGV 2.16.0.

### Transcript capture single nuclei RNA long-read sequencing analysis

Long-read single nuclei RNA sequencing (RNAseq) data were basecalled using Guppy (v6.1.2) with the *dna_r9.4.1_450bps_sup_prom.cfg* configuration file. The run FASTQ was then mapped using Minimap2 (v2.26) with splice aware parameters to hg38, subset for *GBA1* +/-1Mb, and then the subset region was turned back into a FASTQ using samtools (v.1.17) bam2fq. This subset FASTQ was then split into a FASTQ per cell type using corresponding Illumina unique cell type barcodes. Each barcode was matched against the FASTQ allowing for one mismatch and the resulting FASTQs were sorted to only keep unique read ids. Each cell FASTQ was quality controlled using Pychopper (v2.7.1) with standard parameters and mapped using Minimap2 (v2.26) with splice aware parameters. Only reads with a minimum mapping quality of 40 and flagged as primary alignment were kept. Then, we performed the same depth calculations as above using samtools (v1.17) and used the number of cells in the original 10x library as the normalization factor for our depth per region across cell types. Regional *GBA1* and transcript coverage was plotted using ggplot2. As an additional QC, we checked barcode sequence diversity in each of our cell types by calculating the ratio of barcodes in our data divided by total Illumina barcodes. Density plots of barcode usage were generated with ggplot2.

### Illumina short-read sequencing data generation and processing

Illumina short-read data was generated for the same LCLs to complement the ONT data. RNA was extracted in the same method described above, and library preparation and sequencing was completed by Psomagen Inc (https://www.psomagen.com/). The rRNA removed total was fragmented and primed for cDNA synthesis via TruSeq Stranded Total RNA Library Prep kit reagents (96 Samples, Illumina #20020597), and incubated for 8 minutes at 95°C (C1000 Touch Thermal Cycler). The cleaved and primed RNA was reverse transcribed into first strand cDNA using SuperScript II Reverse Transcriptase (#18064-014, Thermo Fisher Scientific, Waltham, MA, USA); Actinomycin D and FSA (First Strand Synthesis Act D Mix) were added to enhance strand specificity. The second strand was synthesized using the 2nd strand master mix from the same TruSeq Stranded Total RNA kit (16°C incubation for 1 hour). To enable adapter ligation, the double-stranded cDNA (dscDNA) was adenylated at the 3’ end, and RNA adapters were subsequently ligated to the dA-tailed dscDNA. Finally, additional amplification steps were carried out to enrich the library material. The final library was validated (D1000 ScreenTape System) and quantified (Quant-iT PicoGreen dsDNA Assay Kit).

The sequencing library was then loaded onto a flow cell containing surface-bound oligos complimentary to the adapters in the library and amplified into distinct clusters. Following cluster generation, the Illumina Sequencing by Synthesis (SBS) technology (Illumina Novaseq 1.5 5000/6000 S4 Reagent Kit-300 cycles [#20028312]; Illumina Novaseq 1.5 Xp 4-Lane Kit [#20043131]) was used to accurately sequence each base pair. Real Time Analysis software (RTA v3) was used to base-call data from raw images generated by the Illumina SBS technology. The binary BCL/cBCL files were then converted to FASTQ files using bcl2fastq (bcl2fastq v2.20.0.422)–an Illumina provided package. Illumina FASTQ data were aligned to hg38 using STAR (v2.7.10) ^28^. We then calculated regional depth following the same steps as the bulk RNA ONT long-read data.

### Accessing publicly available Illumina based whole genome sequencing and RNA sequencing

We additionally looked into Illumina transcriptomic data from the AMP-PD (https://www.amp-pd.org/) and 1000 Genomes (https://www.internationalgenome.org/) datasets for homozygous reference, heterozygous and alternative allele carriers of the rs3115534 variant. For AMP-PD, we extracted data for 1 homozygous G carrier, 47 heterozygous G carriers, and 98 non-European controls. Ancestry of extracted data was divided as follows: 12 African Admixed (7 GT, 5 TT), 15 African (1 GG, 5 GT, 9 TT), 118 European (35 GT, 83 TT), and 1 Asian (1 TT).

For details on AMP-PD samples used see **Supplementary Table 5**. For full details on data generation and processing of WGS and RNAseq see Craig et al. and Iwaki et al. 2021 ^29,30^. For full details on ancestry predictions see Rizig et al. 2023 ^11^. Using these files, we calculated regional depth following the same steps as the bulk RNA ONT long-read data. 1000 Genomes (1000G) WGS and RNAseq data were downloaded and samples were demultiplexed to individual sample FASTQ files and were aligned to hg38 using STAR version (v2.6.1) ^28^. We extracted data for 7 homozygous G carriers, 40 heterozygous G carriers, and 41 homozygous T carriers, all of African ancestry. For details on 1000 Genomes samples used see **Supplementary Table 6**. We calculated regional depth following the same steps as the bulk RNA ONT long-read data. Additionally, we accessed African American ancestry Illumina RNA sequencing data from the Human Brain Collection Core (HBCC) (n=92). Within the 92 samples, 6 were GGs, 20 GTs, and 66 TTs. This data was accessed through https://nda.nih.gov/. FASTQ files were aligned to hg38 using STAR version (v2.6.1) ^28^. For details on Illumina HBCC samples used see **Supplementary Table 7**.

In order to calculate the significance of the rs3115534-G allele on the depth per region, we ran linear regressions in R (v4.3.0) with genotypes as the predictor and normalized depth as the outcome for each dataset and region. Due to low numbers of the GG groups, we combined GG/GT genotypes for the regression in each dataset (Illumina Coriell, ONT Coriell, ONT HBCC, ONT CRISPR, AMP-PD), except for 1000G and Illumina HBCC since there were > 5 samples with a GG genotype. For 1000G and Illumina HBCC we ran the regression with GG/GT/TT split into three groups.

### Validation of the intronic expression GBA1 transcript

To validate the presence of the potential novel *GBA1* intron-containing transcript we generated custom primers that bind to the highest expressed part of the transcript. The forward (GBA1_X11_F9, 5’-GCGACGCCACAGGTAG-’3) and reverse primers (GBA1_X11_R9, 5’-CTTTGTCCTTACCCTAGAACCTC-’3) specific to start before exon 9 and end at exon 11 of *GBA1* were designed and used at a final concentration of 0.8 µM (Integrated DNA Technologies, San Diego, CA)–an additional reverse primer specific to the 3’-untranslated region of *GBA1* (GBA1_UTR_R4, 5’-CCTTTGTCCTTACCCTAGAACC-’3) was also used in conjunction with GBA1_X11_F9 (**Supplementary Table 8**). As input, we used RNA from LCLs with and without the rs3115534 G allele. RNA was quantified using Qubit RNA HS assay (Invitrogen, Q32852) and reverse transcribed to cDNA. cDNA was synthesized from RNA using a high-capacity cDNA reverse transcription kit (Thermo Fisher Scientific, 4368814) following manufacturer recommendations. cDNA then underwent PCR using REDTaq® ReadyMix™ (Millipore Sigma, R2523-20RXN). PCR products were mixed with 6X loading dye (New England Biolabs, B7024S), loaded onto 1% agarose gel containing SYBR™ Safe DNA Gel Stain (Thermo Fisher Scientific, S33102), sized with 1 kb plus DNA ladder (New England Biolabs, N3200L), and imaged on a ChemiDoc Imaging System (Bio-Rad, 12003153). PCR bands were excised from 1% agarose gel and DNA was purified using the NucleoSpin Gel and PCR Clean-Up kit (Takara Bio, 740609) according to manufacturer instructions. Sanger sequencing was performed by Psomagen using conventional protocols (**Supplementary Table 9**)

In addition, we also aimed to assess whether the novel *GBA1* transcript is capped. To check the presence of a 5’ cap, we selected three Coriell lines (ND01137-GG, ND02892-GT, ND22789-TT) for CAGE library prep and sequencing performed by DNAFORM (https://www.dnaform.jp/en/). In brief, RNA quality was assessed with a Bioanalyzer (Agilent) and all samples had a RIN above 8.3. First-strand cDNAs were transcribed to the 5′-ends of capped RNAs and attached to CAGE “bar code” tags. The samples were then sequenced on an Illumina NextSeq 500 and the sequenced CAGE tags were mapped to the human hg38 genome using the BWA software (v0.5.9). Mapped bam files were inspected for transcription start site clusters using IGV (v2.16.0).

### CRISPR editing of rs3115534

To determine whether rs3115534 is the functional variant in this GWAS locus, CRISPR editing was performed by Synthego (https://www.synthego.com/). CRISPR editing was performed using two LCLs ND01137-GG and ND22789-TT with the aim to edit both LCLs to the opposite genotype (**Supplementary Table 10**). In brief, cell pools were created using high-quality chemically modified synthetic sgRNA and SpCas9 transfected as RNPs to ensure high editing efficiencies and without the use of any selection markers that could negatively affect cell biology. Knock-ins were generated using either ssDNA or plasmid, depending on the insert size. The parental cells were electroporated with SpCas9 and target-specific sgRNA to generate the edited cell pool. Similarly, mock transfected cell pools were made by electroporating the parental cells with SpCas9 only and confirmed to be unedited at target locus. After editing, cells (mock transfected, intermediate pools and final fully edited LCLs) were processed for subsequent assays. A predesigned genotyping assay specific to rs3115534 (Thermo Fisher Scientific, C 57592022_20) was used to confirm the CRISPR edited genotypes via an allelic discrimination qPCR assay (QuantStudio 6 Pro, Applied Biosystems, A43159). Additionally, the CRISPR lines were manually inspected post long-read sequencing on IGV (v2.16.0) to confirm no accidental editing of *GBAP1*.

### Bioinformatic annotation of rs3115534

In order to assess the potential functional effect of rs3115534 we investigated several annotation resources. Summary statistics from the largest African PD GWAS were used to generate a LocusZoom plot using AFR linkage disequilibrium patterns ^11^ ^31^. RegulomeDB (v2.2) was explored for rs3115534 to assess its effect on motifs and genome accessibility ^32^. The UCSC Genome Browser was accessed to investigate the conservation of this allele across vertebrates. To evaluate if rs3115534 was involved in splicing we assessed the following algorithms: AGAIN, SpliceAI and Branchpointer ^33–35^. Branchpointer was used in R (v4.3.0) to evaluate the impact of rs3115534 on branchpoint architecture. We ran rs3115534 as the query file and calculated branchpoint predictions using the queryType=SNP option. Gencode’s hg38 v44 release was used as our reference files. Branchpoint predictions were plotted through Branchpointer’s plotBranchpointWindow script. In addition, we assessed all coding and non-coding *GBA1* variants (n=1625) present in gnomAD (v4) for their potential to disrupt intronic branchpoint sequences using AGAIN.

### Assessing the potential protein coding ability of the novel transcript

#### Protein Extraction

LCLs from Coriell Biorepository were maintained in suspension with RPMI 1640 medium (Thermo Fisher Scientific, 11875093) containing 2 mmol/L GlutaMAX (Thermo Fisher Scientific, 35050061), and 15% fetal bovine serum (Thermo Fisher Scientific, A5256701) at 37°C in 5% carbon dioxide. Protein was extracted from LCLs (5e6 cells) via a Tris-HCl cell lysis buffer (Cell Signaling Technology, #9803) containing a Protease/Phosphatase Inhibitor Cocktail (Cell Signaling Technology, #5872) on ice.

#### Multiplex Western Blotting

Protein was normalized to 30 ug and loaded into a 4-20% precast polyacrylamide gel (Bio-Rad, 4561094) before being transferred to a nitrocellulose membrane (Bio-Rad, 1704270). The membrane was blocked for 1 hour (LiCor Biosciences, 927-60001) and incubated (4°C) with primary glucocerebrosidase (Sigma-Aldrich, G4171) and β-actin (Abcam, mAbcam 8224) antibodies on a shaker overnight. Finally, the membrane was incubated with anti-rabbit (LiCor Biosciences, 926-68073) and anti-mouse (LiCor Biosciences, 926-32212) secondary antibodies at room temperature for 1 hour before imaging (Odyssey DLx, LiCor Biosciences). The entire procedure was repeated with identical results.

#### Enhanced Chemiluminescence Western Blotting

Protein was normalized to 30 ug and loaded into a 4-20% precast polyacrylamide gel (Bio-Rad, 4561094) before being transferred to a nitrocellulose membrane (Bio-Rad, 1704270). The membrane was blocked for 1 hour with 5% Blotting Grade Blocker (Bio-Rad, 1706404) and incubated (4°C) with primary glucocerebrosidase (Sigma-Aldrich, G4171) antibody on a shaker overnight in 5% blotting grade block. After washing with TBS-T (0.1% Tween-20), the membrane was incubated with Goat anti-Rabbit IgG (H+L) Cross-Adsorbed Secondary Antibody, HRP (Invitrogen, 31462) for 1 hour. Following washing with TBS-T, the membrane was incubated with anti-β-actin (Abcam, mAbcam 8224) for 1 hour, probed with Goat anti-Mouse IgG (H+L) Cross-Adsorbed Secondary Antibody, HRP (Invitrogen, 31432). The membrane was then probed with Clarity Max Western ECL substrate for 1 hour (Bio-Rad, 1705062). The blot was imaged on the ChemiDoc MP Imaging System (Bio-Rad, 12003153).

#### Mass Spectrometry Analysis of Western Blot

Protein extraction, normalization, and gel-electrophoresis were performed as detailed above. The bands were visualized with Coomassie Blue stain (Bio-Rad, 1610786) and manually excised. Gel bands between 2-15KDa were excised, reduced with 5 mM tris(2-carboxyethyl)phosphine (TCEP) (Sigma Aldrich, 580560), and alkylated with 5 mM N-Ethylmaleimide (NEM), (Sigma Aldrich, 04259). Samples were digested with trypsin (Promega, V5280) at 1:20 (w/w) trypsin:sample ratio at 37°C for 18 hours. Peptides were extracted then desalted using Oasis HLB plate (Waters, WAT058951). LC-MS/MS data acquisition was performed on an Orbitrap Lumos mass spectrometer (Thermo Scientific) coupled to an Ultimate 3000 HPLC. Peptides were separated on a ES902 Easy-Spray column (75-μm inner diameter, 25 cm length, 3 μm C18 beads; Thermo Scientific). Mobile phase B was increased from 3 to 20% in 39 min. Lumos was operated in data-dependent mode. Peptides were fragmented with a higher-energy collisional dissociation method at a fixed collision energy of 35. The Proteome Discoverer 2.4 Software was used for database search using the Mascot search engine. Data was searched against the Sprot Human database.

#### Assessing GBA1 Protein Expression in UK Biobank

In order to replicate previous pQTL results of rs3115534, we accessed the UK Biobank 500K whole genome sequencing data through the UK Biobank Research Analysis Platform (UKB-RAP) (https://ukbiobank.dnanexus.com/). We utilized the population-level variant data produced using Illumina DRAGEN v3.7.8. Genotypes for rs3114435 were extracted using Plink (v.2.0) ^36^ and only those individuals with African or African Admixed ancestry were kept. We then merged the genotype information with available Olink proteomic measures to get a GBA1 protein level per individual. Any NAs from the protein information were dropped. We then generated a violin plot of *GBA1* protein expression per genotype using ggplot2 ^26^ in R (v4.3.0). We ran a linear regression in R (v4.3.0) with genotypes as the predictor and *GBA1* protein levels as the outcome to test for significance.

### Assessing GCase activity in dried blood spots across *GBA1* genotypes

GCase activity was assessed across *GBA1* genotypes in 710 samples containing, 97 non-PD (used as controls), 122 idiopathic PD without a known *GBA1* mutation, 95 *LRRK2* G2019S carriers, 99 rs3115534-GT, 9 rs3115534-GG, and 99 PD risk variant carriers (E326K or T369M), 90 *GBA1* N370S (mild), and 99 *GBA1* severe mutations (e.g. *GBA1* L444P). The glucocerebrosidase enzyme was extracted from one dried blood spot punch (Ø 3.2 mm) per sample and measured by incubating for one hour at 37°C under agitation with an aqueous buffer containing citrate, phosphate, taurocholic acid sodium salt, NaN_3_ and Triton X-100 (pH 5.2). Next, an aqueous solution with the synthetic substrate 4-Methylumbelliferyl ß-D-glucopyranoside and NaN_3_ was added followed by a second incubation step for 16 hours at 37°C under agitation. The enzymatic reaction was quenched by addition of stop buffer (aqueous glycine solution, pH 10.5 adjusted by NaOH). The enzymatic product, 4-Methylumbelliferone (4MU) was quantified by fluorimetry on a microplate reader (Victor X2, PerkinElmer). The instrument was calibrated using an external calibration curve. The enzymatic activity is specified in units of μmol/L/h (amount of product per blood volume per incubation time). Each sample was measured in duplicate and a third replicate was used for background correction. The background of the chemical blank was determined via addition of stop buffer prior to the substrate. As quality parameters for the assay, standard blood samples were added to each batch to ensure the accuracy of the determination.

## Results

### Functional dissection of the GBA1 Parkinson’s disease African ancestry risk locus

Recently, a non-coding *GBA1* variant was reported to be associated with increased risk for PD in African ancestry individuals (**Figure 1A**) ^11^. This variant was also reported to be an eQTL and pQTL resulting in increased gene expression (**Figure 1B**) and decreased protein expression (**Figure 1C**) ^12,13^. Using UK Biobank Olink data, we replicated the previously reported association between rs3115534 and *GBA1* protein levels. After filtering for African ancestry, a total of 1147 samples remained with a *GBA1* protein measure (43 GG, 351 GT, 753 TT). The G allele is significantly associated with lower *GBA1* protein levels (p=0.006). (Supplementary Figure 1).

**Figure 1:**
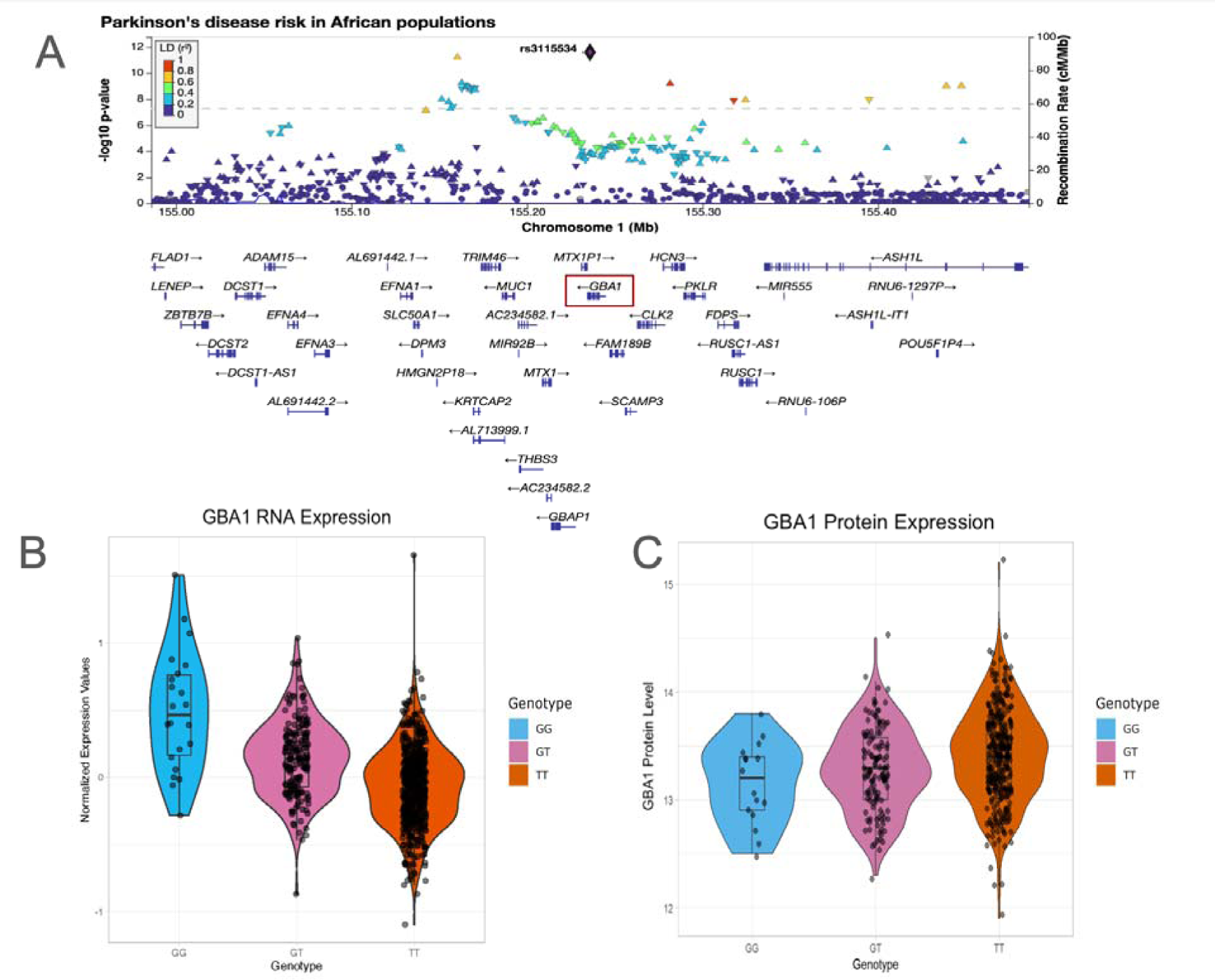
Overview of the African ancestry Parkinson’s disease *GBA1* GWAS locus. A) LocusZoom plot showing rs3115534 as index variant (purple diamond) and located in intron 8 of *GBA1*. B) rs3115534 as eQTL for *GBA1* RNA expression from Kachuri et al 2023 showing increased GBA1 expression with G risk genotypes. C) rs3115534 as pQTL for *GBA1* protein expression from Surapaneni et al 2022 showing decreased protein levels with G risk genotypes. For all boxplots, the center line represents the median, edges of box represent Q1 and Q3, and ends of bars represent the maximum and minimum not including outliers.

Given the complexity of short read DNA and RNA mapping in the *GBA1* region due to high sequence homology with *GBAP1*, we generated ONT long-read RNAseq data to investigate this region in a more accurate and comprehensive manner. Long-read RNAseq data was generated from eight African ancestry LCLs from Coriell across risk genotypes (1 homozygous risk-GG, 4 heterozygous risk-GT, 3 homozygous non risk-TT). Surprisingly, we identified that there was a clear enrichment of sequence reads in the intron 8 region proximal to exon 9 that was specific to carriers of the G risk allele (**Figure 2A, Supplementary Table 11).**

**Figure 2:**
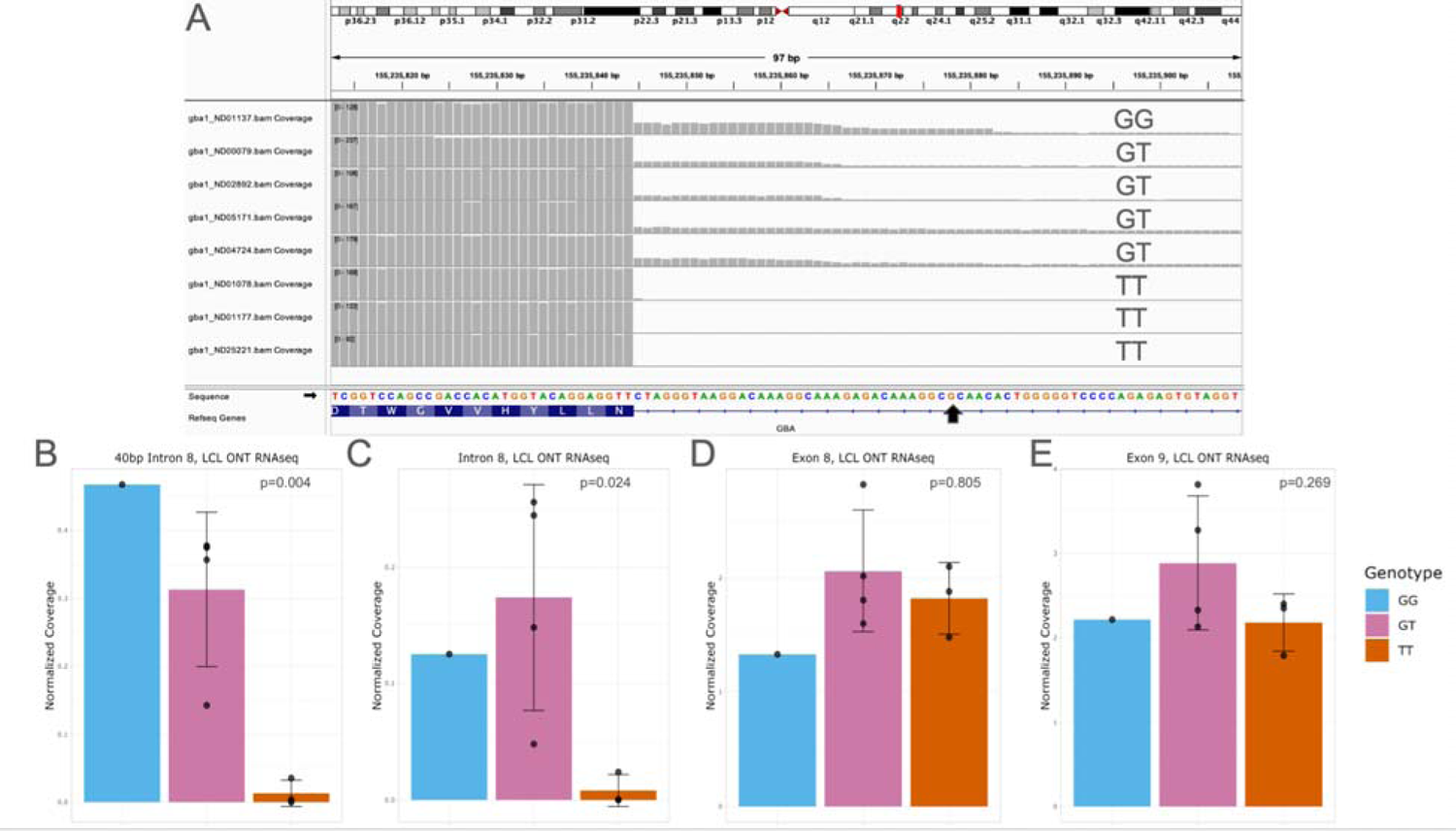
*GBA1* intron 8 expression is correlated with rs3115534 genotype. **A)** Oxford Nanopore Technologies long-read RNA sequencing of 8 lymphoblastoid cell lines (LCL) shows a consistent pattern where the rs3115534-G risk allele is associated with intron 8 expression and absent in the homozygous T (non-risk allele) individuals generated using Integrative Genomics Viewer. **B)** Quantification of intron 8 expression is significantly associated with the G allele in both the 40bp region prior to exon 9 and **C)** the full intron 8 (linear regression, p < 0.05). **D,E)** No significant differences were identified in the two neighboring exons 8 and 9. Coverage for all panels normalized by dividing mean depth by total number of mapped reads per million as detailed in methods. Error bars represent standard deviation for all panels.

Subsequently, *de novo* StringTie2 isoform calling suggested there were multiple novel transcripts including one starting approximately 40 base pairs before exon 9, and two full length *GBA1* isoforms including all of intron 8, hereafter collectively named as “intron 8 expression” (Supplementary Figure 2). Interestingly, the reported index variant rs3115534 is included in intron 8 expression (**Figure 2A**, Black arrow). We quantified the expression levels of this region and determined that expression levels of intron 8 were highly correlated with the presence of the G genotype (**Figure 2A,B,C, Supplementary Table 12**). In addition, no clear differences were observed for neighboring exons 8 and 9 (**Figure 2D,E**) nor in total *GBA1* or *GBAP1* expression (Supplementary Figure 3). Importantly, every mapped sequence read exhibits a G allele, indicating their origin from the G risk haplotype. We manually changed the base in G reads to T to confirm that these reads map uniquely to *GBA1* and that the striking difference we observed was not an artifact of mismapping to *GBAP1* driven by the rs3115534 variant. No differences in mapping were observed (Supplementary Figure 4).

Next, we examined whether intron 8 is also expressed in the human brain. Using ONT long-read RNAseq of 8 African ancestry HBCC frontal cortex samples from different genotypes (4 GG, 2 GT, 2 TT), we ran StringTie2 with the same methods as the LCLs. However, even when using our custom transcript models with our transcripts of interest, StringTie2 only identified the shorter intron 8 transcript in one GG carrier, likely due to lower expression of *GBA1* in the brain and lower RNA quality, as evidenced by lower RIN values (**Supplementary Table 1, Supplementary Table 12**). However, when looking at expression coverage plots, intron 8 expression was observed in all carriers of the G allele (GG, GT) and not in TT carriers (**Figure 3A**, Supplementary Figure 5**, Supplementary Table 11**). Next, we generated Illumina short read RNAseq for 18 LCLs including the initial 8 from above and identified a similar, though less pronounced, unexpectedly increased read coverage across intron 8 (**Figure 3B, Supplementary** Figure 6). Likewise, similar enrichment in the intron 8 region all correlated with the G allele of rs3115534 was seen in Illumina RNAseq data from HBCC frontal cortex (**Figure 3C, Supplementary** Figure 7), 1000 Genomes LCL RNAseq (**Figure 3D**) and AMP-PD blood based RNAseq data (**Figure 3E**), importantly when increase numbers a clear allelic dosage effect becomes visible i.e., GG has significantly more intron 8 expression compared to GT carriers (Supplementary Figure 8 and 9**, Supplementary Table 11**).

**Figure 3:**
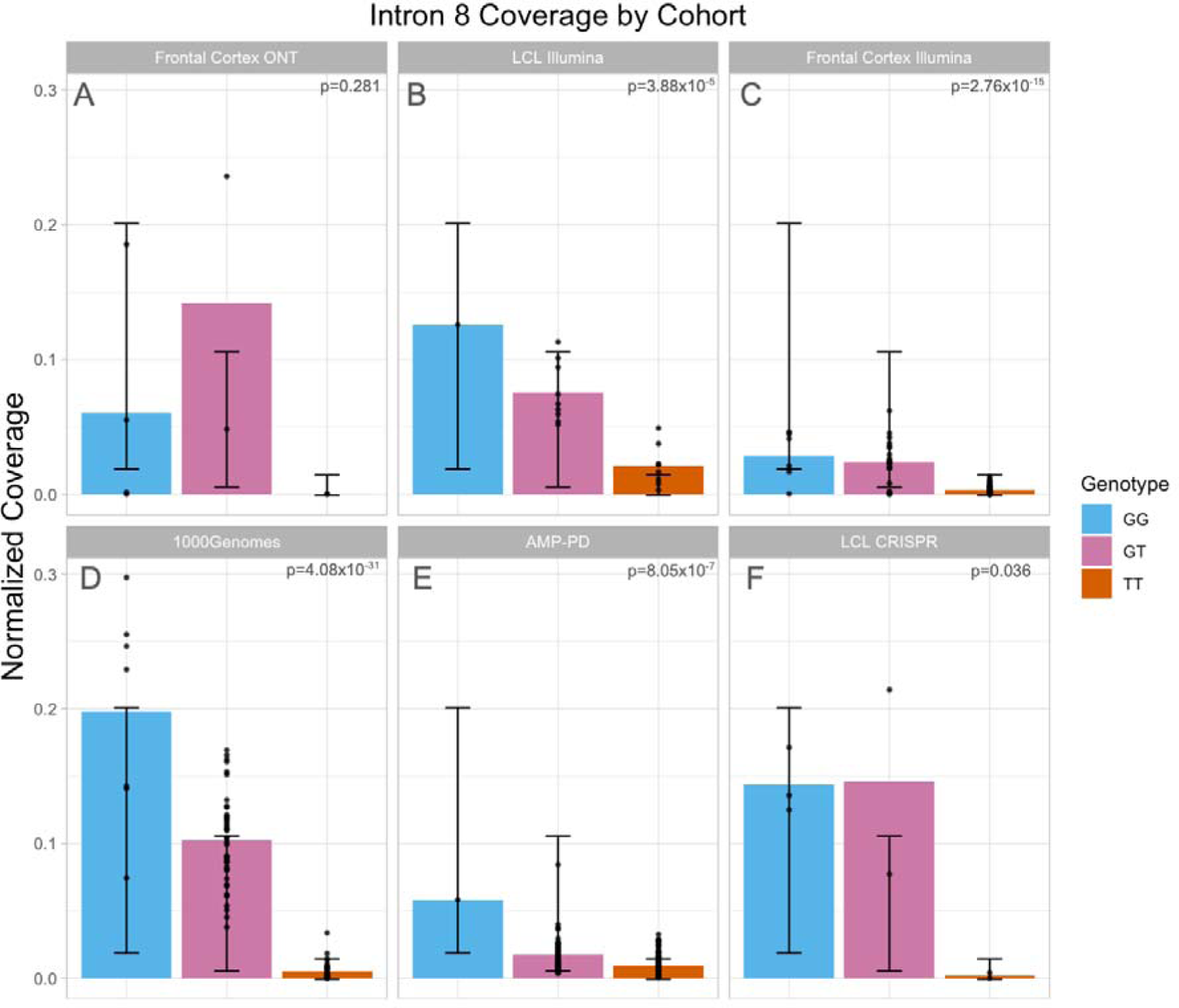
Increased intron 8 expression across datasets in G allele carriers. **A)** Intron 8 coverage from human frontal cortex sequenced with Oxford Nanopore (n=8). Intron 8 expression is only present in G allele carriers but does not reach statistical significance (p=0.281) likely due to a smaller sample size. **B)** Intron 8 coverage from LCLs (n-18) sequenced with Illumina. Expression is significantly associated with G allele (p=3.88E-5). **C)** Intron 8 expression from human frontal cortex sequenced with Illumina (n=92). Expression is significantly associated with G allele (p=2.76E-15). **D)** Intron 8 coverage from LCLs in 1000 Genomes cohort (n=88). Expression is significantly associated with G allele (p=4.08E-31). **E)** Intron 8 coverage from blood in AMP-PD cohort (n=148). Expression is significantly associated with G allele (p=8.05E-7). **E)** CRISPR editing of two lymphoblastoid cell lines shows that the rs3115534-G risk allele is significantly associated with intron 8 expression (p=0.036). Coverage for all panels normalized by dividing mean regional depth by total number of mapped reads per million as detailed in methods. Error bars represent standard deviation for all panels.

### Risk allele specific GBA1 intron expression does likely not code for a new protein product

Due to the comparatively lower *GBA1* expression in frontal cortex tissues and the greater accessibility of LCLs, the majority of subsequence experiments and analyses were conducted using LCLs. To validate the inclusion of *GBA1* intron 8, we designed primers specific to the most highly expressed region proximal to exon 9. Reverse transcriptase PCR (RT-PCR) validated the presence of this novel intron 8 containing transcript in LCLs only in G risk variant carriers, confirming our initial findings (Supplementary Figure 10 and 11). Next, we used CAGEseq, a technique often used to accurately pinpoint transcription start sites. The CAGEseq library preparation is dependent on the presence of an RNA cap, which is used to capture RNAs. We detected the canonical *GBA1* transcription start site, however we did not identify a unique transcription start site within intron 8 (Supplementary Figure 12). This indicates that the intron 8-containing transcript does not have a 5’ cap and likely starts at the canonical transcription start site of *GBA1*. However, the novel transcripts do have a poly-A tail that can be detected with ONT long-read RNAseq.

The protein coding capacity of this transcript was assessed via multiplexed and enhanced chemiluminescence (ECL) western blotting. Given that the highest intronic expression was observed close to exon 9, an C-terminus antibody specific to amino acids 517-536 of the functional GCase protein was selected to ensure the capture of proteins that have a more downstream start site. Both multiplex and ECL methods consistently identified the native GCase protein but showed no other bands, indicating a lack of novel protein (Supplementary Figure 13). Subsequent proteomic analysis of the region that was predicted to harbor the protein of the novel transcript did not identify any known *GBA1* peptides (**Supplementary Table 13**). These results suggest that the transcript is not protein coding and that the disease mechanism is likely to be RNA-based.

### rs3115534 is the functional risk variant at the GBA1 locus

The close proximity of the index risk variant to the novel transcript and the lack of other variants in linkage disequilibrium nearby makes rs3115534 a strong candidate to be the functional effect variant for disease risk (**Figure 1A**). To investigate this, we CRISPR-edited two LCLs: one in which the homozygous risk genotype (GG) was edited to be homozygous non-risk (TT), and one in which the homozygous non risk (TT) was edited to be homozygous risk (GG) (**Supplementary Table 10**). Genotyping qPCR was used to confirm the rs3115534 genotype in the edited lines. All contained the genotype as expected except one, in which full conversion from GG to TT was only partially successful and led to a heterozygous GT line. This line was excluded from downstream analysis.

Next, we performed ONT long-read DNA and RNAseq to confirm successful CRISPR editing and to assess the presence or absence of the novel intron 8 expression. ONT long-read DNA showed successful editing of TT to GG and partial editing of line ND01137 from GG to GT, validating the qPCR results. ONT long-read RNAseq again showed the presence of intron 8 sequence reads but only in LCLs carrying a G allele (Supplementary Figure 14). Quantification of the intronic reads again displayed an effect specific to the G allele (**Figure 3F**), showing that the rs3115534-G allele is solely responsible for intron 8 transcription in LCLs.

### Intron 8 is expressed across cell types in the human brain

Next, we assessed in which brain cell type this novel transcript is expressed. Initial screening of frontal cortex brain single nuclei RNAseq showed that overall *GBA1* expression is too low to accurately assess transcript expression, and intron 8 expression was not observed (Supplementary Figure 15). Therefore, we used an enrichment strategy with probes targeting the *GBA1* region in the single nuclei cDNA library. After enrichment, we extracted *GBA1* transcripts and performed long-read ONT RNAseq on the single nuclei libraries from 1 GG carrier and 1 TT carrier. Sequencing showed clear enrichment for *GBA1* and *GBAP1* transcripts (**Supplementary Table 14**). *GBA1* and *GBAP1* were ubiquitously expressed across major brain cell types (Supplementary Figure 16). Supporting our previous results, reads covering intron 8 were predominantly identified in the GG carrier as shown in above sections (Supplementary Figure 16). When assessing the expression of the intron 8 region across cell types, the region is ubiquitously expressed across cell types, similar to the full *GBA1* transcript.

### rs3115534 is located in a key branchpoint sequence of intron 8 in GBA1

To investigate potential functional downstream consequences of rs3115534 we explored several *in silico* algorithms. RegulomeDB analysis to assess rs3115534 effect on motifs and genome accessibility reported that this variant is an eQTL and is located in a region of open chromatin. However, no transcription factor motifs are affected by this variant. Sequence conservation analysis showed that the T allele (non-risk) is highly conserved across vertebrates, and humans are the only vertebrates harboring G as a reference allele (Supplementary Figure 17). Interestingly, when analyzing the 5-Methylcytosine (5mC) DNA modifications in the ONT long-read DNA data of CRISPR-edited LCLs, we identified that the G allele is methylated. When rs3115534 is a T, the methylation is lost (Supplementary Figure 18). This change in methylation status was confirmed when ONT sequencing was performed on the initial LCLs (Supplementary Figure 19) and two additional frontal cortical brain samples (Supplementary Figure 20). Finally, given that rs3115534 was close to an exon (34 base pairs), we also assessed if the variant potentially disrupts splicing. To assess potential splicing disruptions we used two complementary approaches 1) SpliceAI, which is based on a deep neural network that accurately predicts splice junctions from pre-mRNA transcript sequences and 2) Branchpointer and AGAIN, which are algorithms that are driven by an understanding of splicing biology. Using SpliceAI, no significant score of interest (<0.2) was identified. However, using Branchpointer and AGAIN, we identified that this variant is located within the key intronic branchpoint sequence of intron 8. The rs3115534-G allele (risk, C in the minus strand) is likely disrupting the splicing process by removing the key A base (non-risk, T in the positive strand) (**Figure 4**).

**Figure 4:**
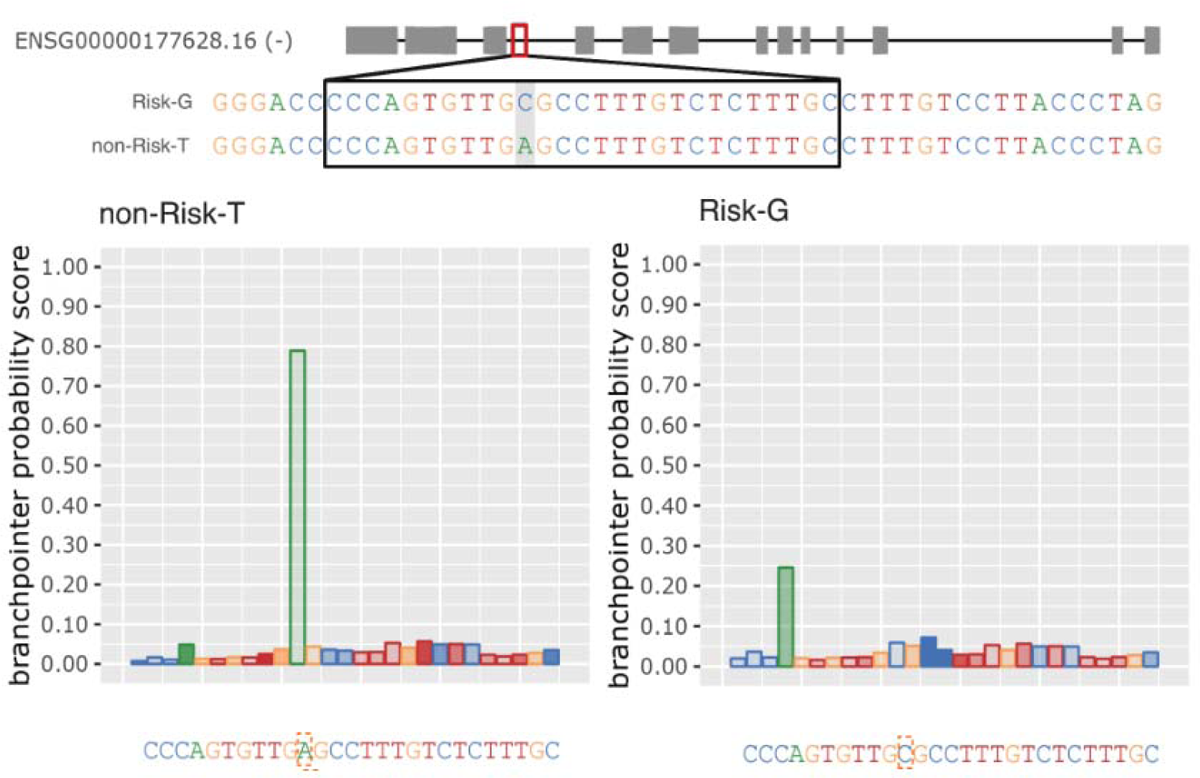
The *GBA1* intronic rs3115534 variant acts as a splicing branchpoint. The causal variant rs3115534 (highlighted in gray on the top and in red dashed box on the bottom) in intron 8 is located in the key splicing branchpoint according to Branchpointer. When rs3115534 in non-risk state (T), on the antisense strand, the A allele functions as a branch site for the spliceosome, while in the risk state (G), on the antisense strand, the C allele disrupts this branch site resulting in abnormal splicing.

This explains our initial finding from the ONT long-read RNAseq, which suggested new transcripts were transcribed from the G risk haplotype (Supplementary Figure 2). The “shorter transcripts”, starting close to exon 9, are spliced at the AG sequence (CT on sense strand) 6 bases upstream of rs3115534 and are sequenced as a novel transcript given they retain the poly A tail. The longer transcripts, containing the full not spliced intron 8, are sequenced in full and also arise exclusively from the G risk haplotype. Here, intron 8 is not spliced out due to the branchpoint disruption. Given these results, we also wanted to assess potential consequences on GCase activity levels. Using Centogene’s CentoCards we compared GCase activity across individuals without PD, idiopathic PD, *GBA1* carriers with PD coding risk variants (E326K, T369M), *GBA1* carriers with mild Gaucher coding risk variants (N370S), *GBA1* carriers with severe Gaucher coding risk variants (including L444P) and rs3115534 variant carriers in heterozygous (G/T) and homozygous state (G/G). A significant dose dependent reduction of GCase activity correlated with the G risk genotype was observed (**Figure 5A**). Notably, the G allele–associated reduction in GCase activity is similar to PD coding risk variants that do not cause Gaucher disease (e.g. E326K) and higher than the Gaucher causing variant, confirming that the reduction of GCase is not enough to cause Gaucher (**Figure 5B**). Combined with our other results, this data suggests that while no new protein product is made, the risk allele disrupts normal splicing and creates a new transcript that is likely non-functional leading to a decrease in overall GCase protein levels and therefore also lower GCase activity (**Figure 6**).

**Figure 5:**
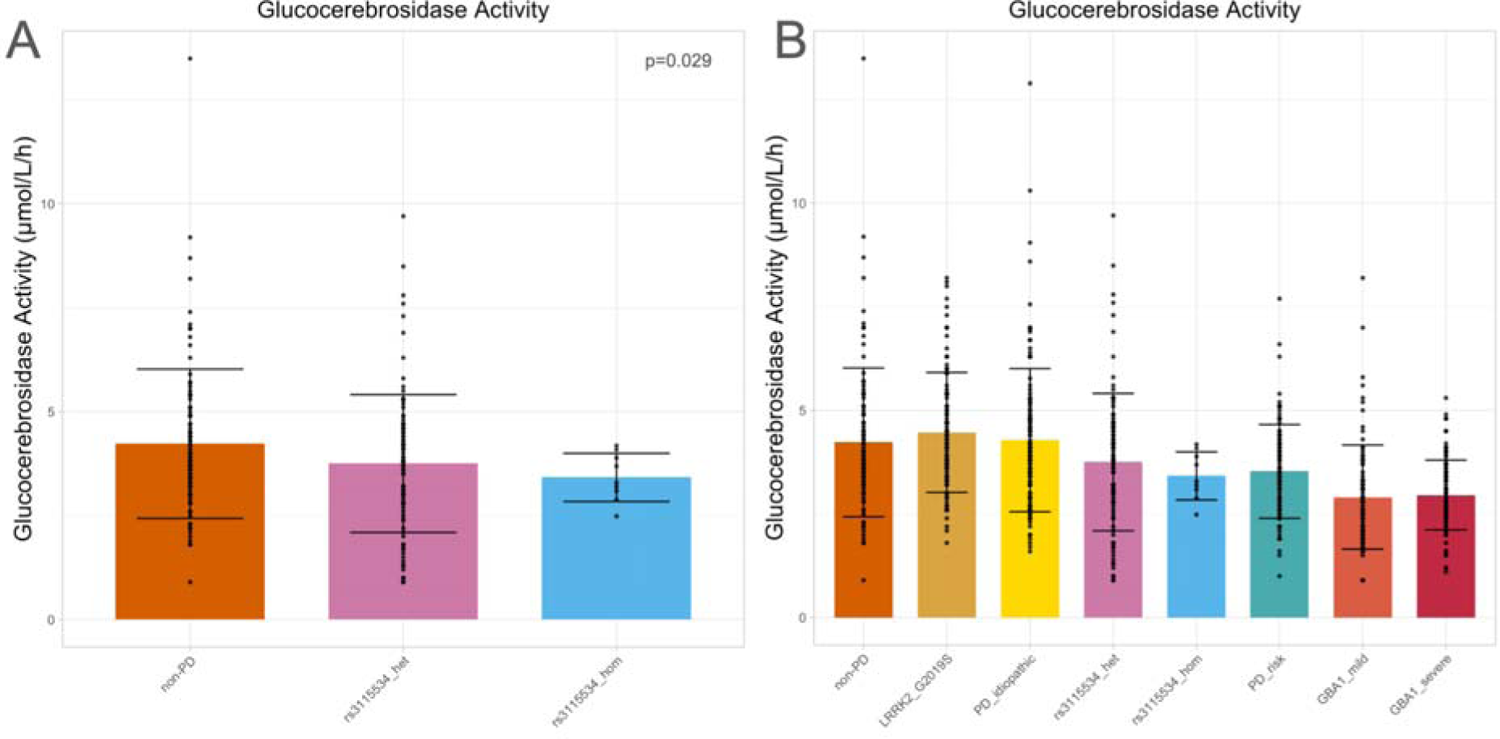
Measuring GCase activity across genotypes. A) GCase activity was measured across rs3115534 *GBA1* genotypes showing a significant dose G-allele dependent reduction across genotypes. B) When measuring GCase activity across multiple *GBA1* genotypes it appears that rs3115534-GT and rs3115534-GG reduce GCase activity to similar levels as PD risk variants (E326K and T369M) but remain higher than *GBA1* Gaucher causing variants (GBA1_mild eg. N370S and GBA1_severe eg. L444P).

**Figure 6:**
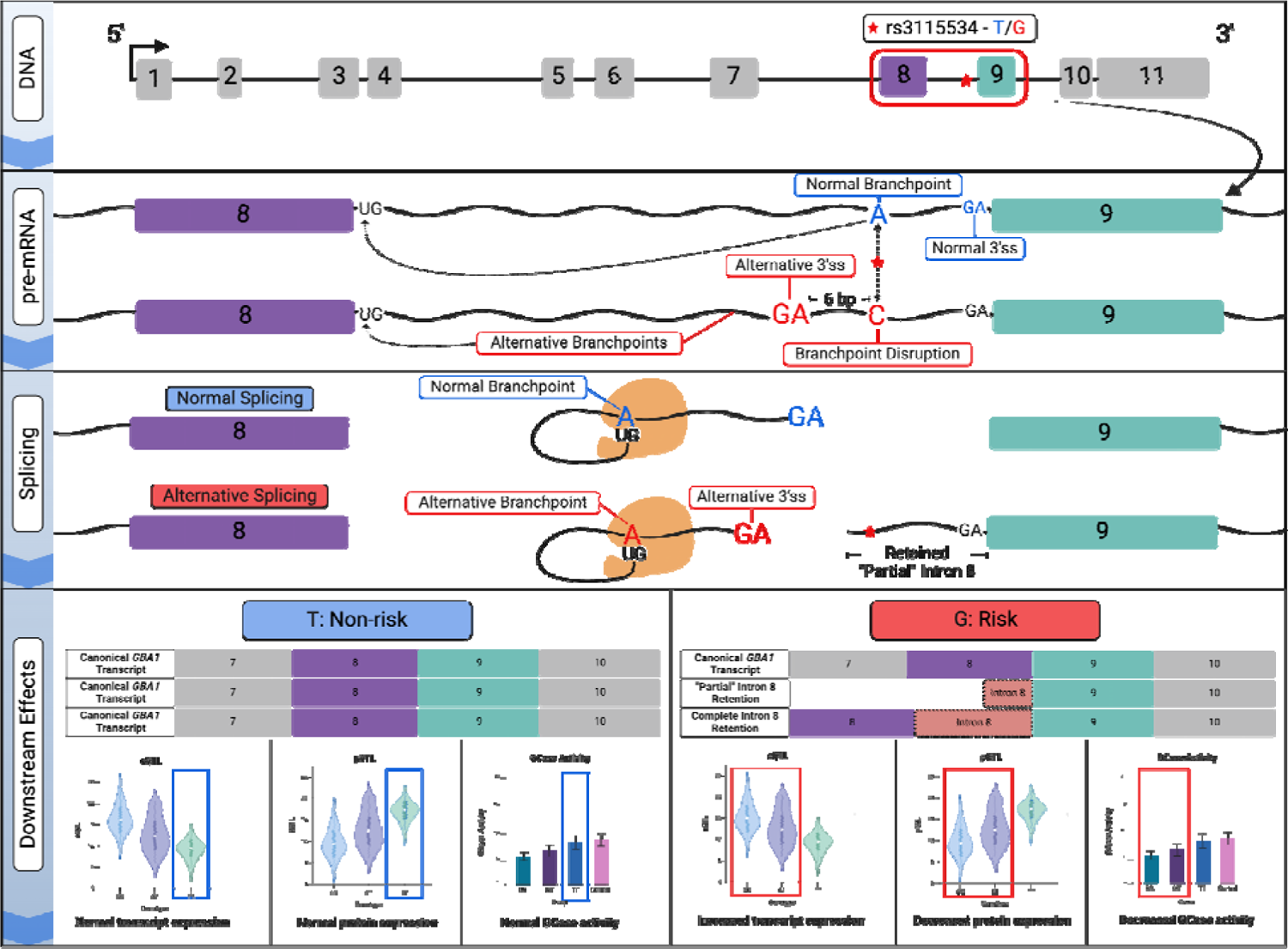
Suggested variant to function hypothesis of rs3115534. rs3115534-T is transcribed to pre-mRNA as an A–a highly conserved branchpoint nucleotide. However, rs3115534-G, which confers elevated PD risk, is instead transcribed to a C, causing the observed branchpoint disruption (rs3115534 denoted as red star). This single nucleotide change in intron 8 impacts splicing by disrupting the normal binding from the adenosine branchpoint nucleotide and the 5’ splice site (5’ss) (GU). Subsequently, an alternative branchpoint is utilized, uncovering an alternative 3’ splice site (3’ss) upstream of the normal splice site immediately proximal to exon 9 and resulting in partial and complete intron retention and fewer functional *GBA1* mRNA transcripts. Downstream, abnormal splicing of *GBA1* leads to reduced GCase protein and subsequent lower GCase activity which is a known pathomechanism of PD/DLB. 3’ and 5’ss refers to splice sites. Generated with biorender.com.

Finally, we wanted to assess if other variants in the *GBA1* region potentially affect branchpoint sequences. When exploring all *GBA1* variants from gnomAD (n=1625), three variants were identified to be of interest by the AGAIN algorithm, including the rs3115534 variant (**Supplementary Table 15**). The two other variants, rs140335079 and rs745734072, did not show a similar Branchpointer pattern (Supplementary Figure 21). When exploring intron expression for rs140335079 in AMP-PD, we did not identify any differences across genotypes (Supplementary Figure 22). No RNAseq data was available for rs745734072 given it’s a rare variant and only identified in the South Asian population.

## Discussion

Most of the current genetic, genomic and functional knowledge in the DLB and PD field is based on European ancestry findings. Recent efforts are showing great progress in making genomics more diverse including GWAS reporting in East Asia, South Asia, Latin America, and Africa ^11,37–39^. In the past decade, GWAS has been the workhorse in genetics and has shown tremendous progress in the identification of genomic regions associated with traits and diseases ^40^. However, one of the main limitations of GWAS is that it cannot pinpoint the exact mechanism, causal gene, or variant in associated genomic regions. Therefore, follow-up methods like QTL analysis and functional studies are used to fill this gap.

Recent progress in African ancestry PD genetics identified a non-coding *GBA1* risk variant associated with disease risk and an earlier age at onset ^11^. Here, we show extensive follow-up of genomic and transcriptomic data from this region, pinpoint the functional variant, and highlight the likely variant-associated mechanism. Using long-read RNA sequencing, we identified intron 8 retention, which was enriched in close proximity to the reported index variant rs3115534. Expression levels of intron 8 were correlated with G allele dosage and very low or absent in TT carriers. Given the high sequence overlap between *GBA1* and *GBAP1*, it is unlikely that this expression is simply a sequence mapping artifact. The absence of a CAGEseq peak in this intronic region and no additional protein isoform suggests that the risk mechanism is RNA-based. CRISPR experiments showed that the reported index variant rs3115534 is the variant responsible for directly changing transcription. Importantly, given the significant evidence for an intronic branchpoint disruption of rs3115534, it is clear that this is the cause of the intron 8 expression.

RNA splicing is the process where pre-mRNA is transformed into mRNA by removing introns. This complex process depends on the donor site (GT), the acceptor site (AG), and the branch site (often a sequence motif including an adenine base). Disruption of any of those sequences causes missplicing, which typically results in reduced functional mRNA. In the case of rs3115534, the A allele (non-risk, T in the sense strand) is part of the key branch site sequence of intron 8 and therefore when rs3115534 is mutated to the C allele (risk, G in the sense strand) partial splicing disruption occurs (**Figure 6**). This disruption results in partial intron 8 retention, this becomes particularly clear when investigating the long-read sequencing data which shows enrichment of sequence reads close to exon 9, and starts by the next intronic AG acceptor site 39 bases prior to exon 9, which is often seen by other branch site sequence disruptions ^41^.

Our finding that branchpoint disruption may confer disease risk highlights a novel mechanism for increased PD risk. All prior knowledge of *GBA1* disease mechanisms has attributed damaging coding variation to reduced GCase activity, leading to disease risk. Here we show that the branchpoint disruption likely causes splicing dysregulation, which results in increased risk by lowering the GCase protein levels (pQTL) and therefore also reducing the GCase activity correlated with genotype dosage. This is consistent with the hypothesis that reduction in *GBA1* activity is the pathomechanism underlying PD/DLB and highlights a novel therapeutic target for African ancestry individuals.

Importantly, there are several inherent limitations of the study, many of which are driven by the lack of ancestrally diverse tissue, cellular, and data resources. First, given the high sequence homology between *GBA1* and *GBAP1* and the low expression levels of the potential novel transcript isoforms, we cannot exclude its protein coding potential. It could be possible that this isoform is too lowly expressed or is degraded too quickly for the antibody to detect the protein expression or to be seen on generalized mass spectrometry analysis. Second, although we measure GCase activity to assess potential downstream effects of the rs3115534 variant and several other *GBA1* genotypes, this assay remains variable across samples. Here we include a large collection of samples and observed expected effects across *GBA1* genotypes and thus believe the results are robust. However, more research is needed in larger collections and across different cell types, including neuronal cell types, to dissect the exact reduction in GCase levels. Ongoing recruitment efforts in the Global Parkinson’s Genetics Program (https://gp2.org/) aim to fill that gap ^42^.

In summary, here we report rapid translation of a novel African ancestry GWAS hit and show the power of genetic diversity, which can result in valuable new biological insights into well-studied genes like *GBA1*. We provide compelling evidence that rs3115534-G is the causal variant and show that the likely functional mechanism is a disruptive allelic change in the intronic branchpoint sequence that disrupts splicing resulting in reduced protein levels. This shows how a common GWAS variant can be functionally explained via an intronic branchpoint sequence alteration, a potentially underexplored mechanism for GWAS. Overall, this implies that rs3115534-G is the most common damaging *GBA1* variant with a minor allele frequency of over 20% in certain populations, and for which a substantial number of West African PD cases where there are heterozygous (40%), or homozygous (13%) risk carriers ^11^. There is no evidence that this variant causes Gaucher disease, a condition that is remarkably infrequent in African ancestry groups; which is likely due to the magnitude of the biological effect of this variant, where the result is only a partial reduction of GBA1 protein levels and GCase activity. Interestingly, this is a novel mechanism for increased disease risk at *GBA1* and no other variants in *GBA1* were identified to act via a similar mechanism. The frequency of this risk variant, its mapping as the functional allele at this risk locus, and the variant’s mode of action at an intronic branchpoint make this an attractive candidate for precision-based therapeutics, in a remarkably underserved population. This work underscores the scientific and societal importance of working in groups underrepresented in research, the need for the generation of biological and data resources in these groups, and marks the start of the realization of the mission of the Global Parkinson’s Genetics Program.

## Supporting information

Ethics Statement

## Supplementary Figures

**Supplementary Figure 1:**
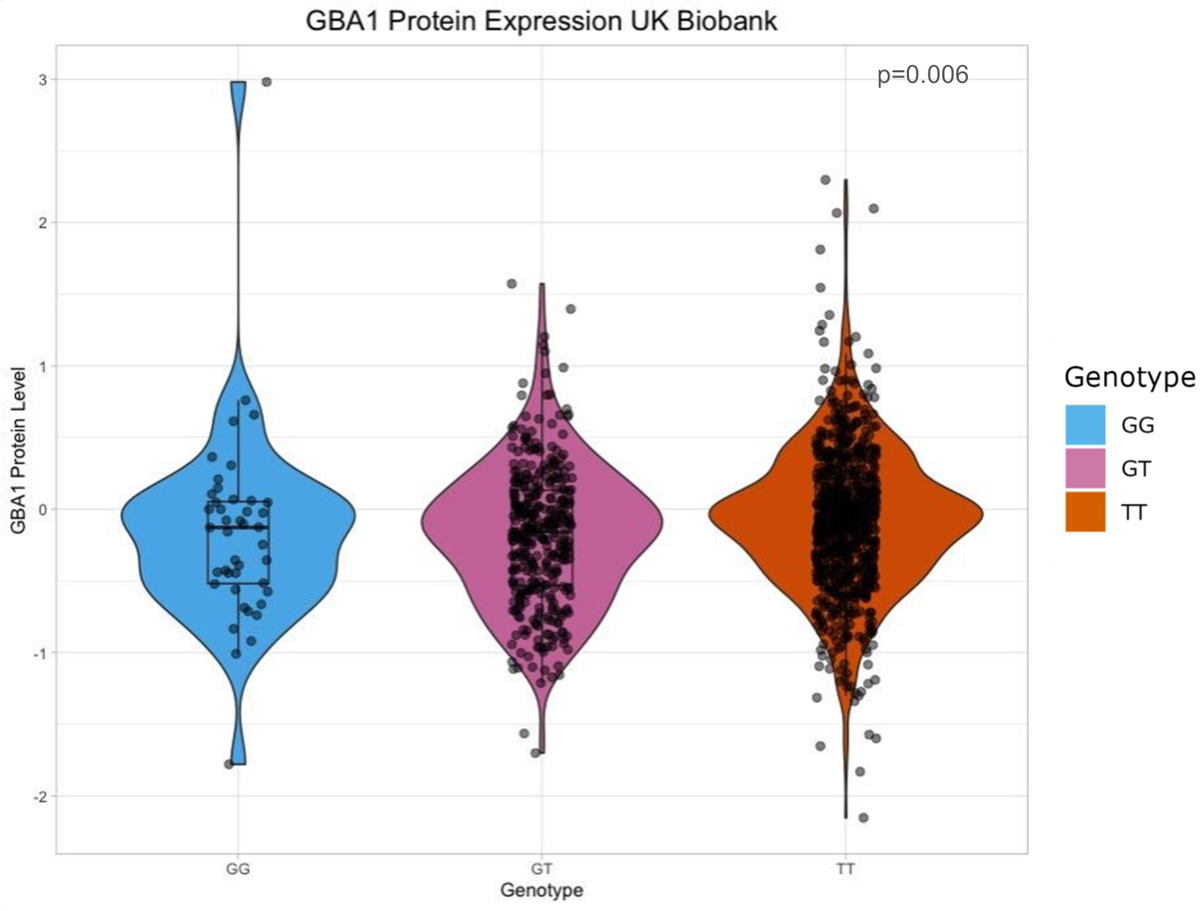
rs3115534 lowers total *GBA1* protein expression in the UK Biobank dataset (n=1147). rs3115534-G significantly lowers total *GBA1* protein expression (p < 0.05) in the UK Biobank dataset. Only African and African-Admixed individuals were kept for this pQTL analysis. This replicates the previously reported pQTLof rs3115534 in the African American population (see Figure 1). The observed decrease in protein levels is further supported by reduced GCase activity levels.

**Supplementary Figure 2:**
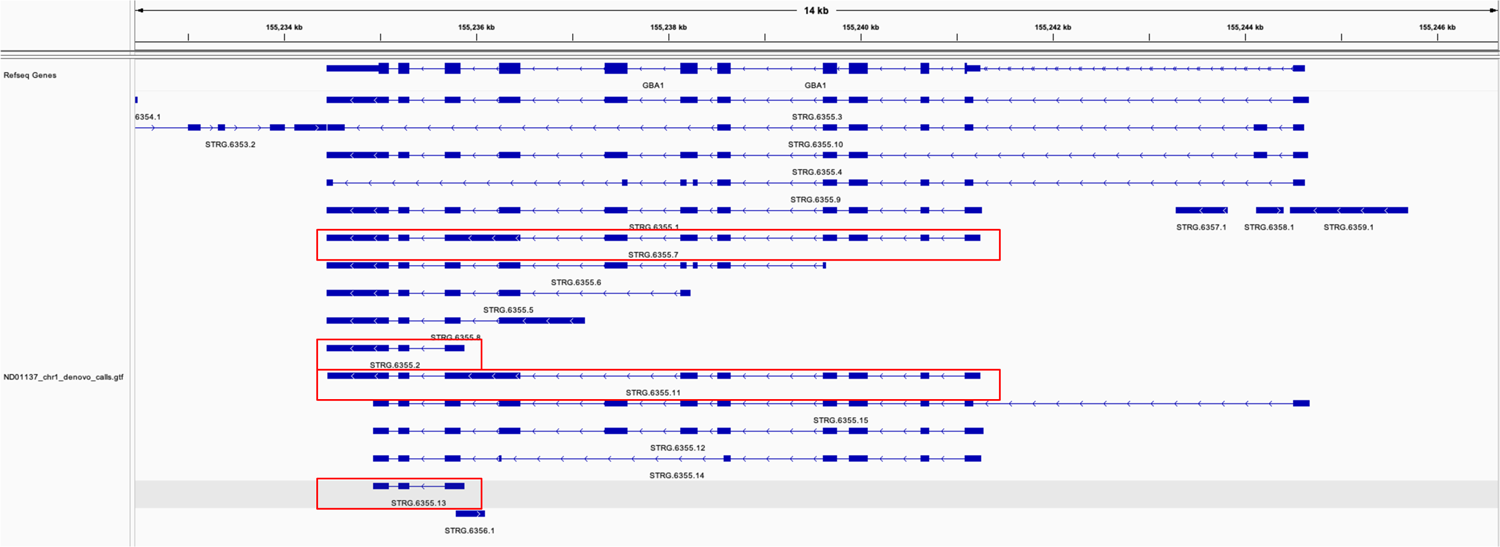
Stringtie2 isoforms from GG sample. Stringtie2 was run *de novo* in long-read mode for ND01137, a rs3115534-GG lymphoblastoid cell line sequenced with long-read RNA sequencing ONT protocols. STRG.6355.2, STRG.6355.7, STRG.6355.11, and STRG.6355.13 in the red boxes show abnormal intron 8 splicing.

**Supplementary Figure 3:**
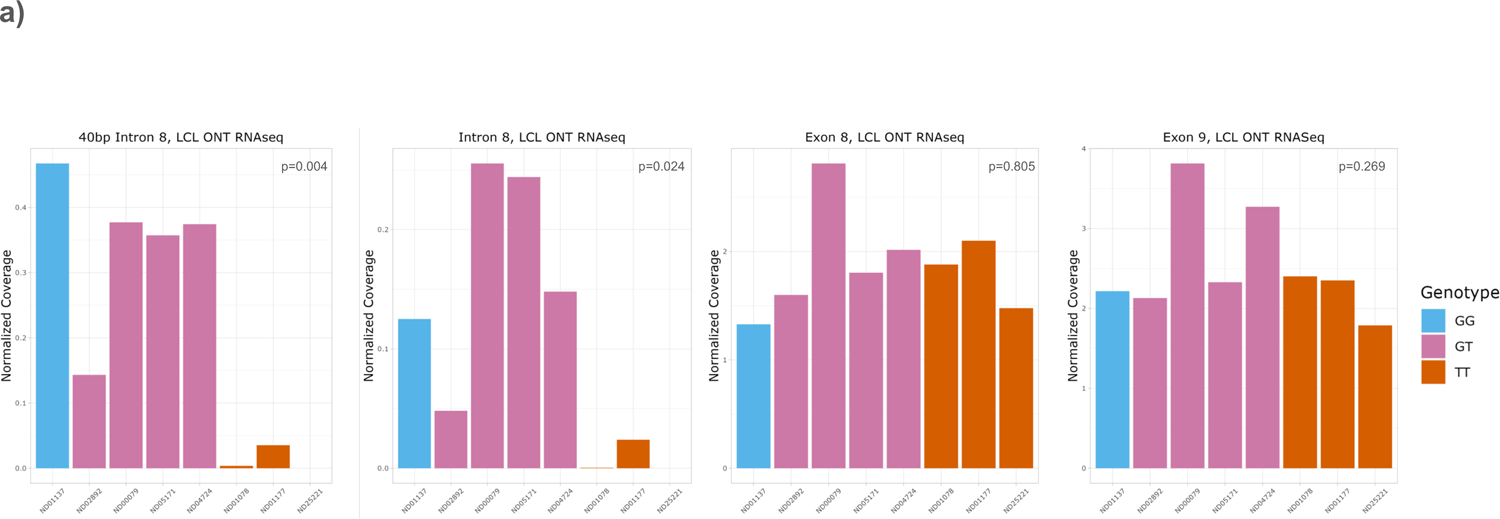
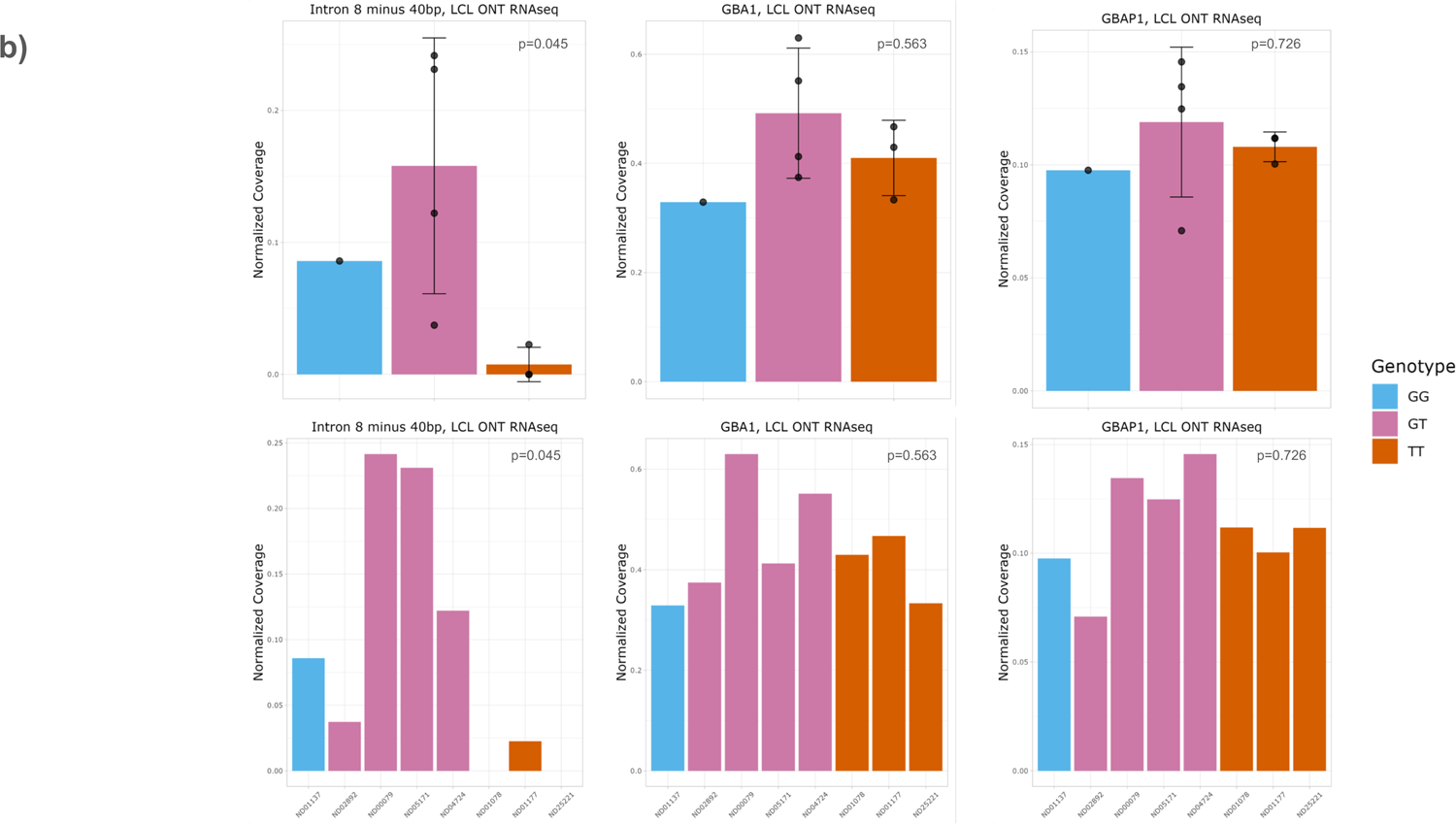
Additional coverage plots for lymphoblastoid cell lines sequenced with Oxford Nanopore Technologies long-read RNA sequencing. a) Per sample quantification of intron 8 expression matching Figure 1 genotype collapsed per genotype plots including 40bp intron 8 region prior to exon 9, full intron 8, exon8 only and exon 9 only. Coverage for all panels normalized by dividing mean depth by total number of mapped reads per million as detailed in methods. b) Additional coverage plots from LCLs for intron 8 minus 40bp transcript region, *GBA1*, and *GBAP1*. Plot are shown both per sample and collapse by genotype. No significant differences shown in G allele carriers *GBA1* and *GBAP1* gene expression. Error bars represent standard deviation for all panels.

**Supplementary Figure 4:**
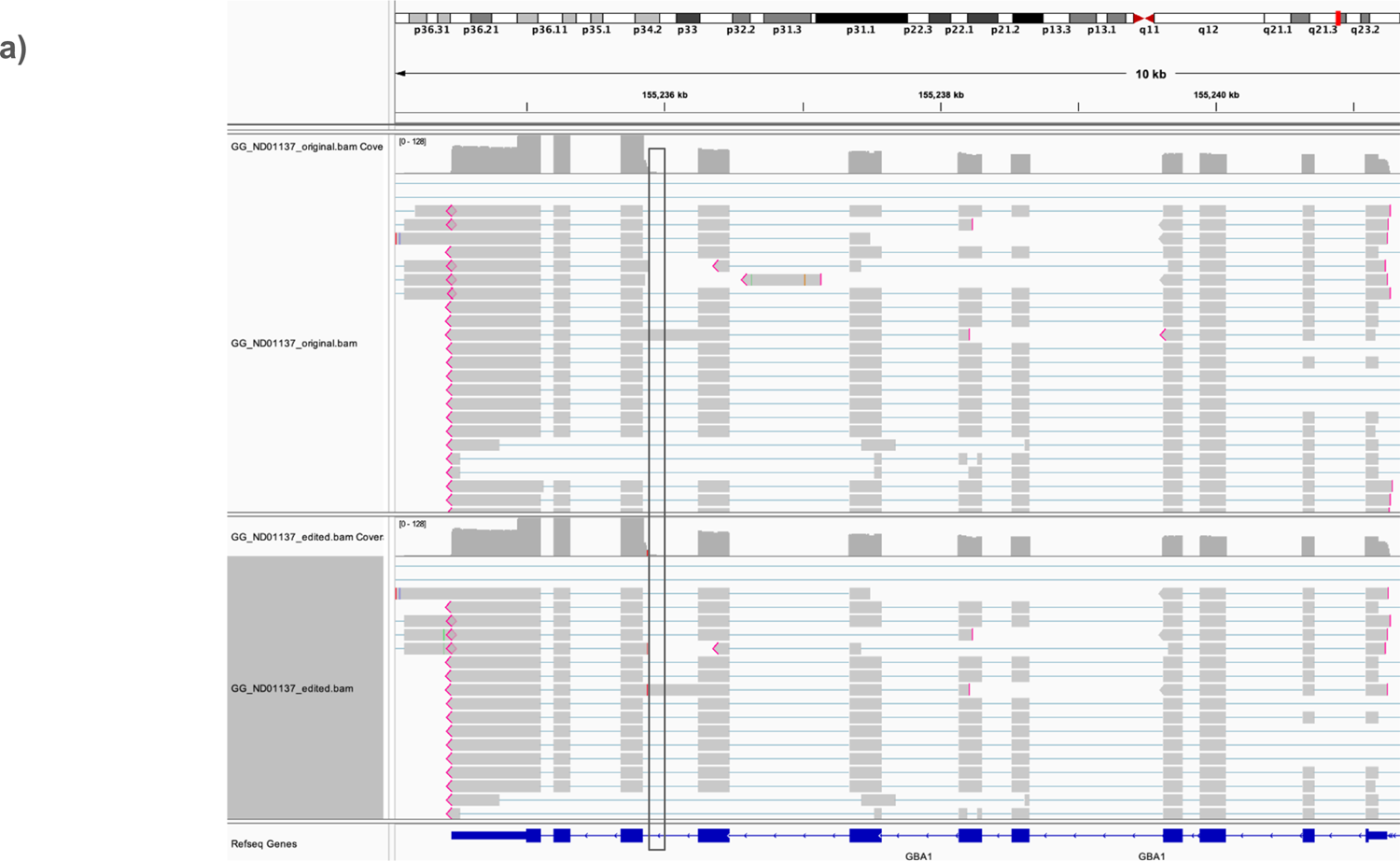
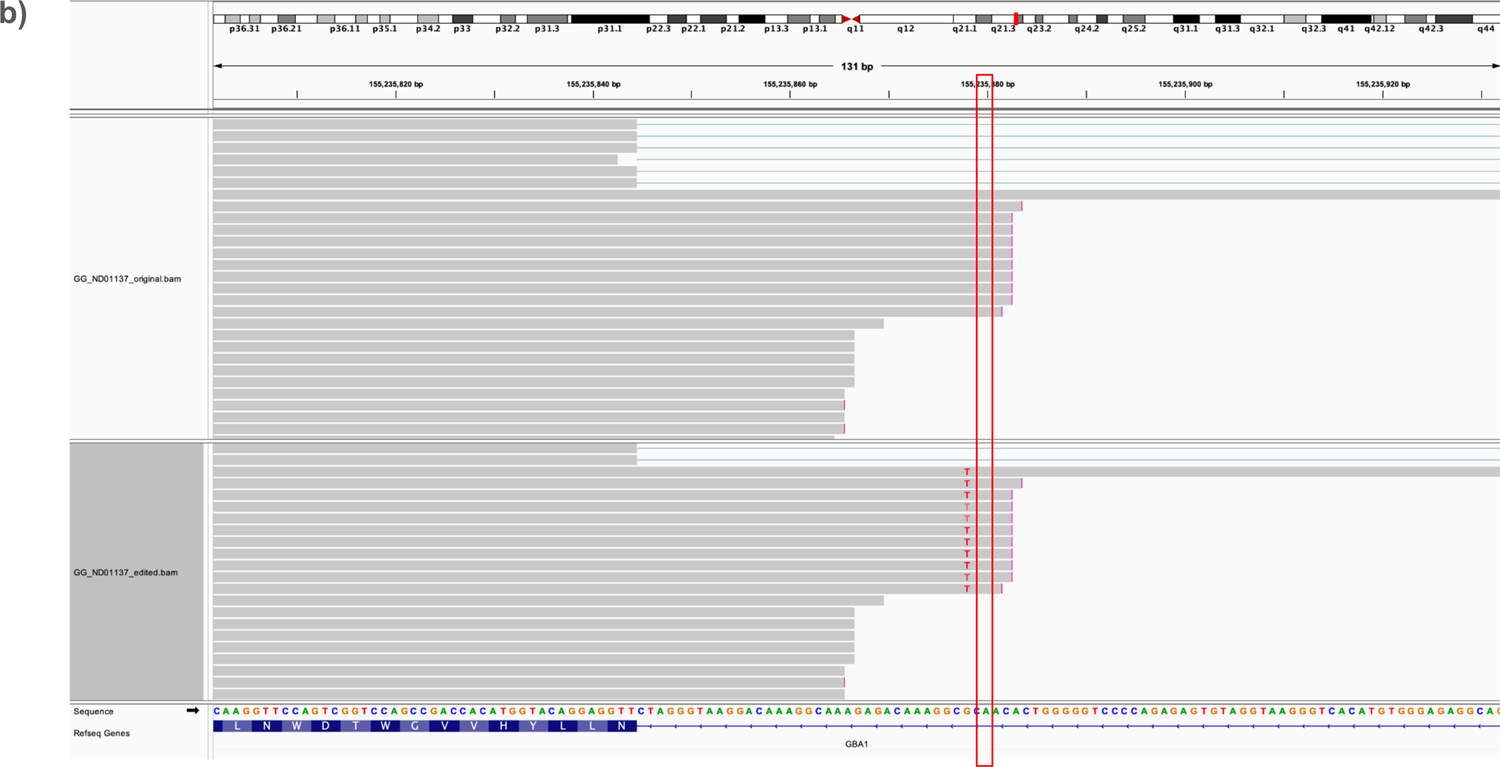
Integrative Genome Browser inspection of Oxford Nanopore Technologies long-read RNA sequencing mapped reads in lymphoblastoid cell line ND01137 rs3115534-GG bam and *in silico* edited to ND01137 rs3115534-TT bam. a) Zoomed out IGV capture of *GBA1* reads in both files, black box denoting intron 8 area of interest. No obvious mapping differences are observed with the manually edited base. b) Identical to a) mapped reads zoomed in on rs3115534 to confirm successful manual *in silico* edit from GG to TT at this locus.

**Supplementary Figure 5:**
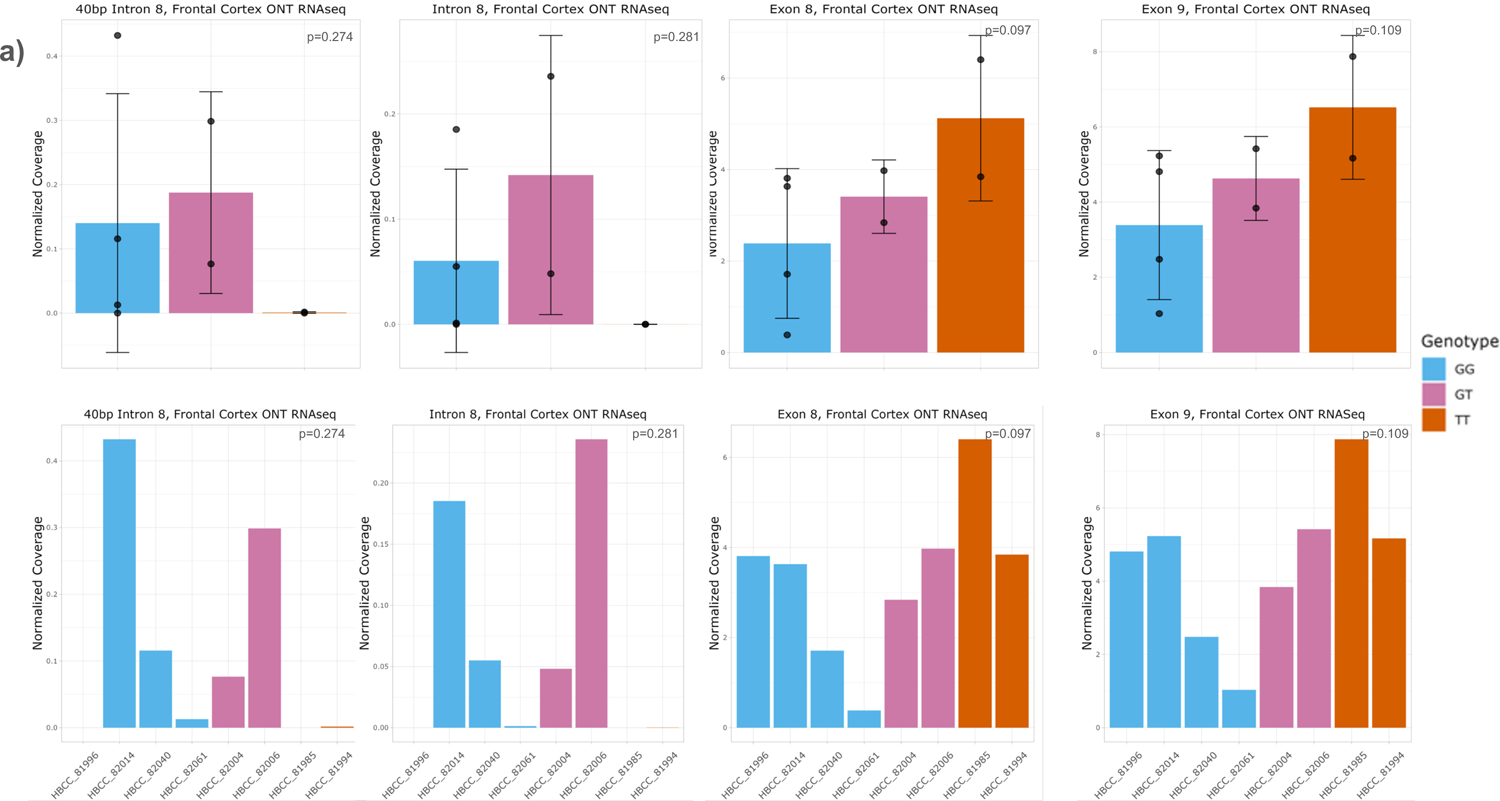
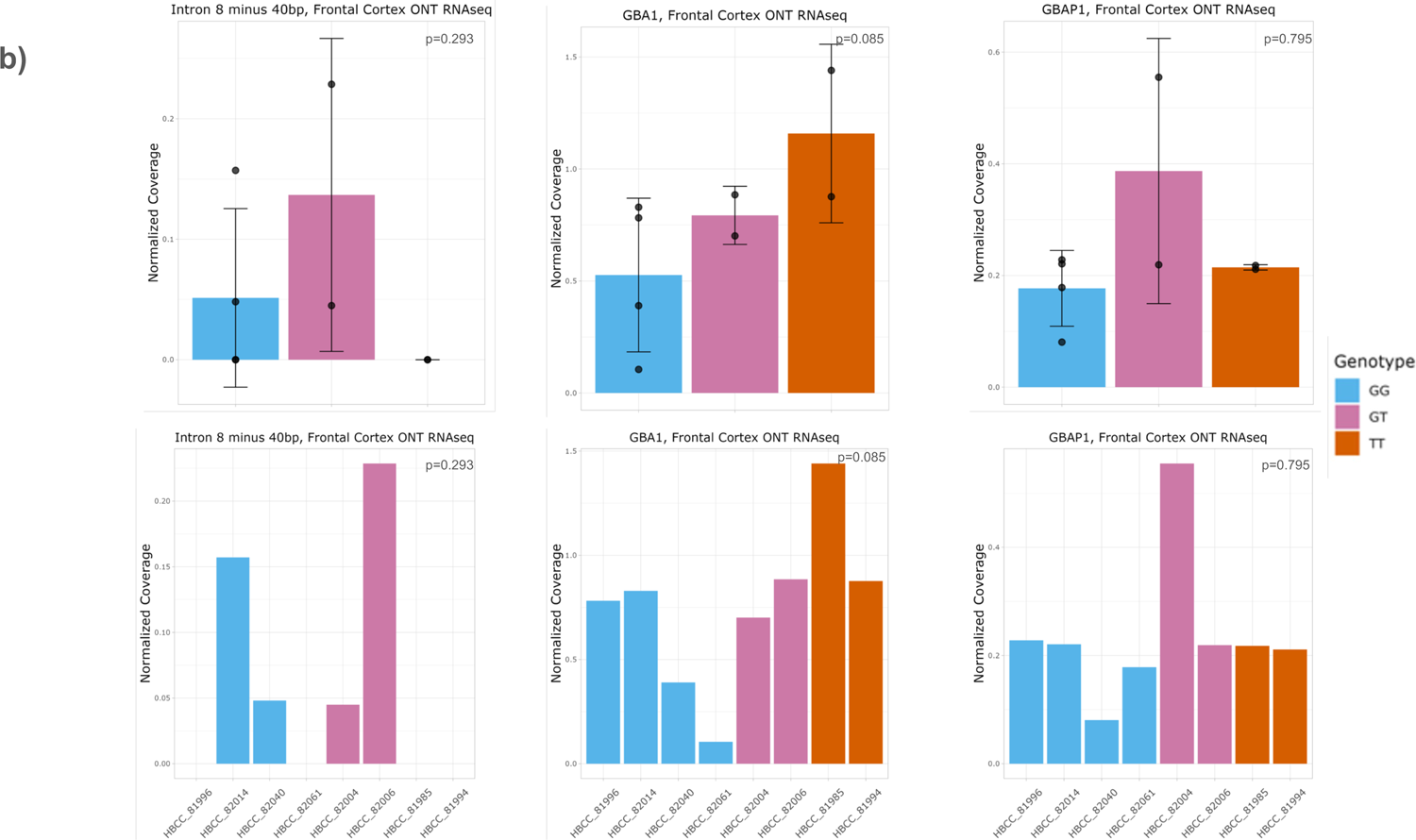
Regional coverage plots from human frontal cortex Oxford Nanopore Technologies long-read RNA sequencing showing enrichment of intron 8 expression in rs3115534-G carriers (n=8). a) Coverage plots of the highly expressed intron 8 40bp region, intron 8, exon 8, and exon 9. Rs3115534-G carriers have a higher expression of intron 8 regions. Plots shown per sample and collapse by genotype. b) Coverage plots of intron 8 minus 40bp region, *GBA1*, and *GBAP1*. Plots shown per sample and collapsed by genotype. Error bars represent standard deviation for all panels.

**Supplementary Figure 6:**
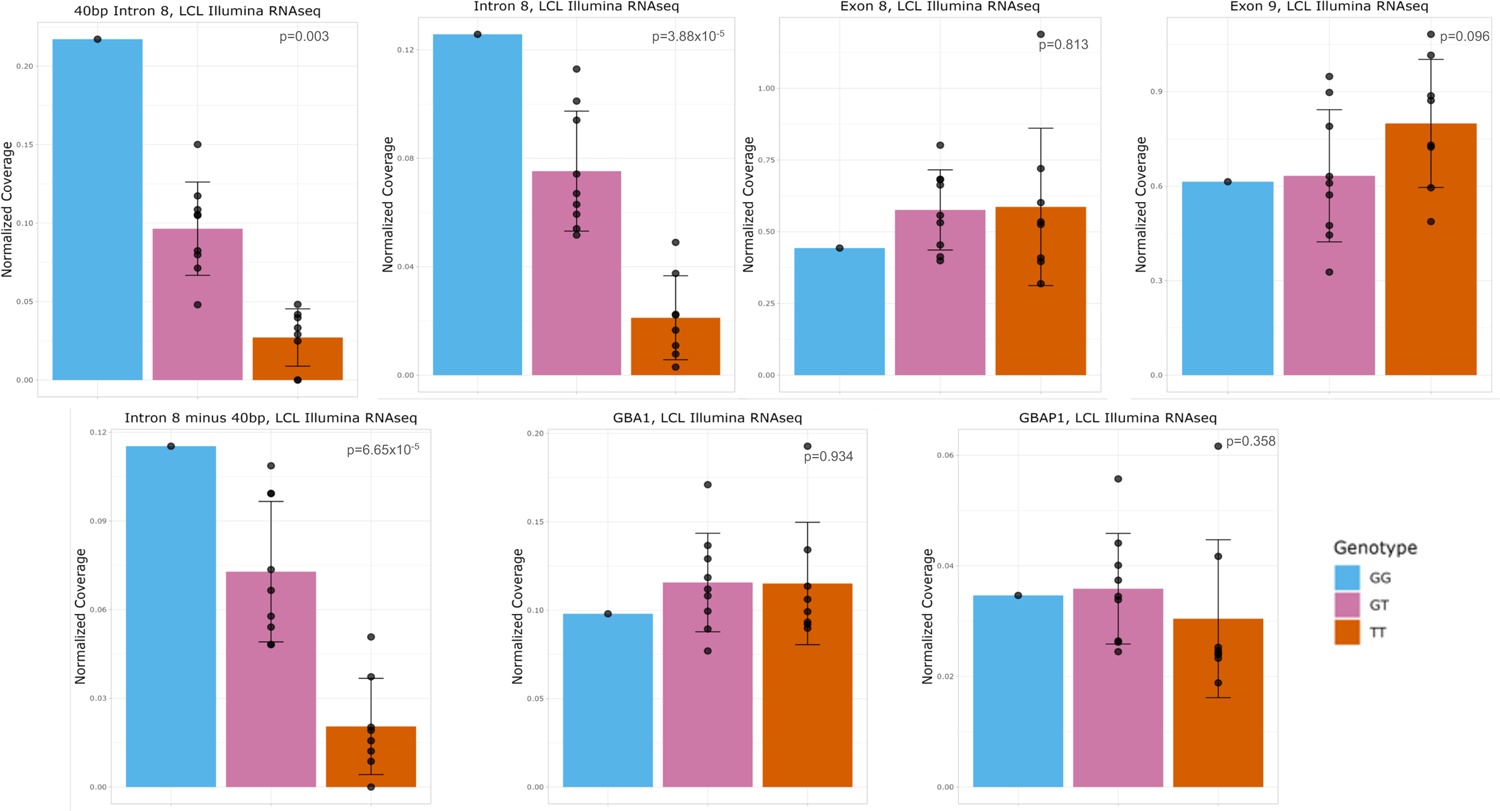
Regional coverage plots from lymphoblastoid cell lines sequenced with Illumina RNA sequencing showing enrichment of intron 8 expression in rs3115534-G carriers (n=18). Coverage plots of regions of interest collapse by genotype from lymphoblastoid cell lines similar as previous supplementary figures. Enrichment of intron 8 expression in G allele carriers still present with short-read sequencing. Error bars represent standard deviation for all panels.

**Supplementary Figure 7:**
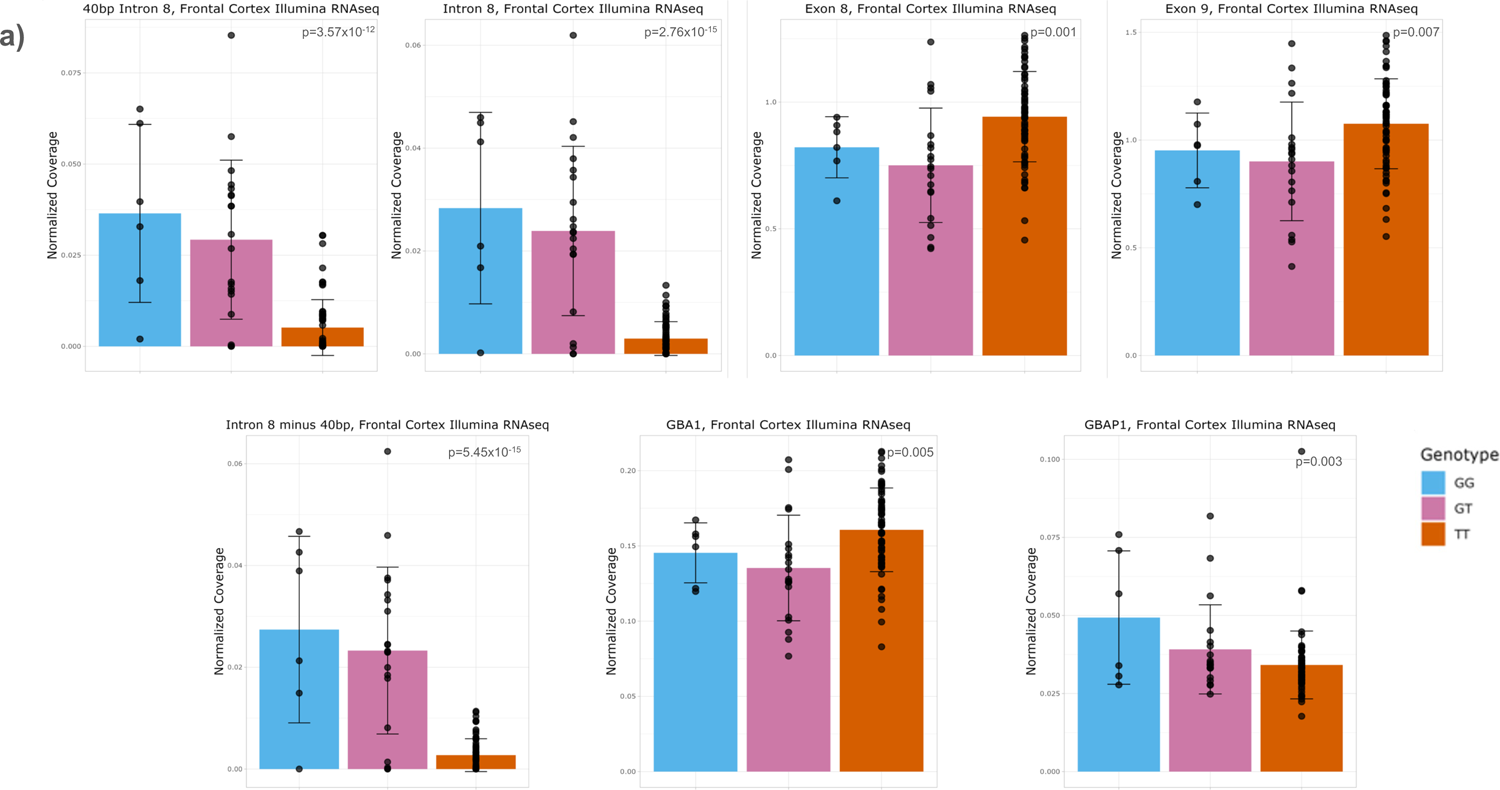
Regional coverage plots from human frontal cortex Illumina short-read RNA sequencing showing enrichment of intron 8 expression in rs3115534-G carriers (n=92). Coverage plots of the highly expressed intron 8 40bp region, intron 8, exon 8, and exon 9, intron 8 minus 40bp region, *GBA1*, and *GBAP1.* rs3115534-G carriers have a higher expression of intron 8 regions. Plots shown collapsed by genotype. Error bars represent standard deviation for all panels.

**Supplementary Figure 8:**
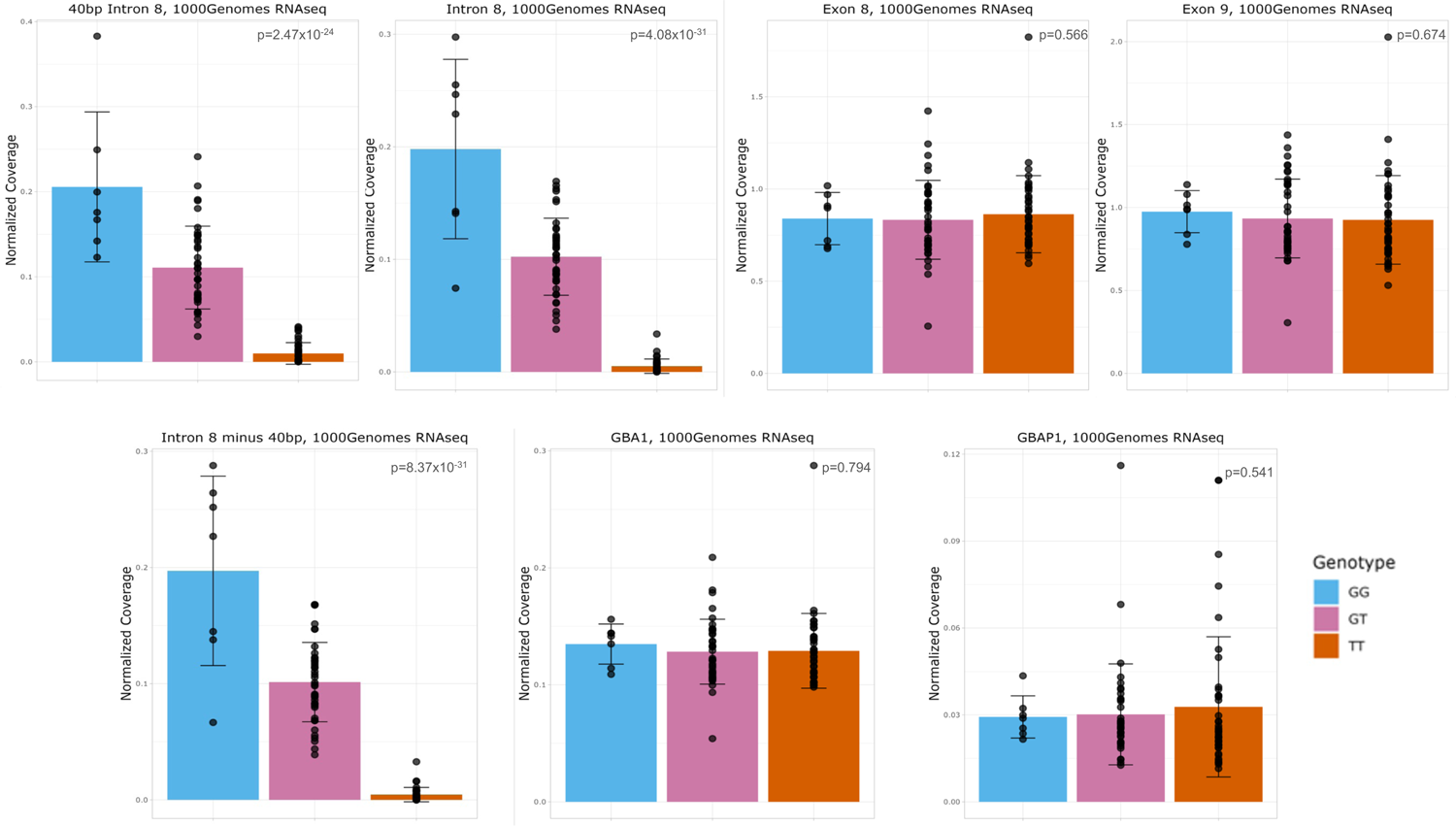
Regional coverage plots from 1000 Genomes dataset showing enrichment of intron 8 expression in rs3115534-G carriers (n=88). Coverage plots of regions of interest collapsed by genotype similar as previous supplementary figures. Enrichment of intron 8 expression in rs3115534-G allele carriers still present with larger sample-sizes. Error bars represent standard deviation for all panels.

**Supplementary Figure 9:**
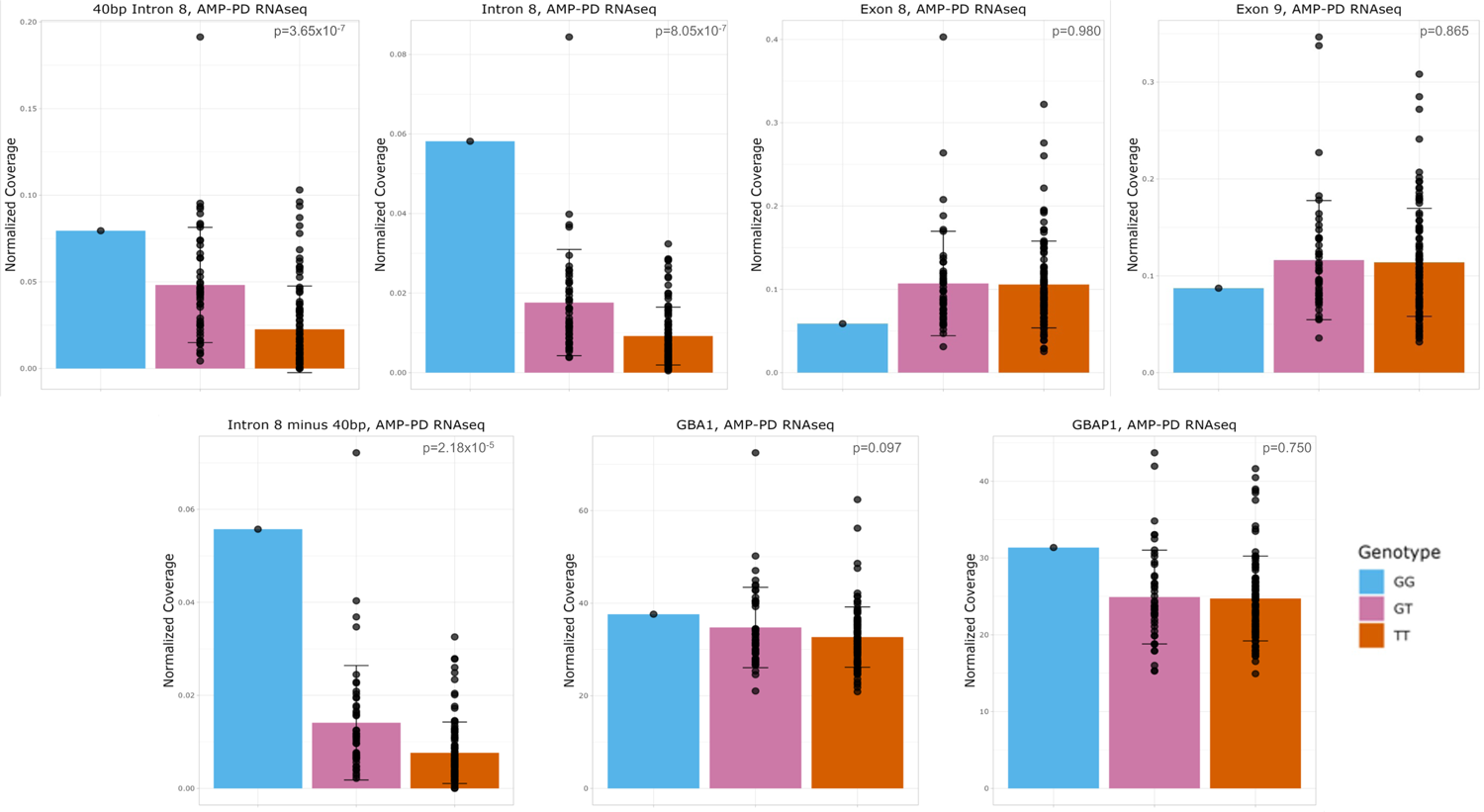
Regional coverage plots from AMP-PD blood-based RNA sequencing showing enrichment of intron 8 expression in rs3115534-G carriers (n=146). Coverage plots of regions of interest collapse by genotype similar as previous supplementary figures. Enrichment of intron 8 expression in rs3115534-G allele carriers still present in blood samples. Error bars represent standard deviation for all panels.

**Supplementary Figure 10:**
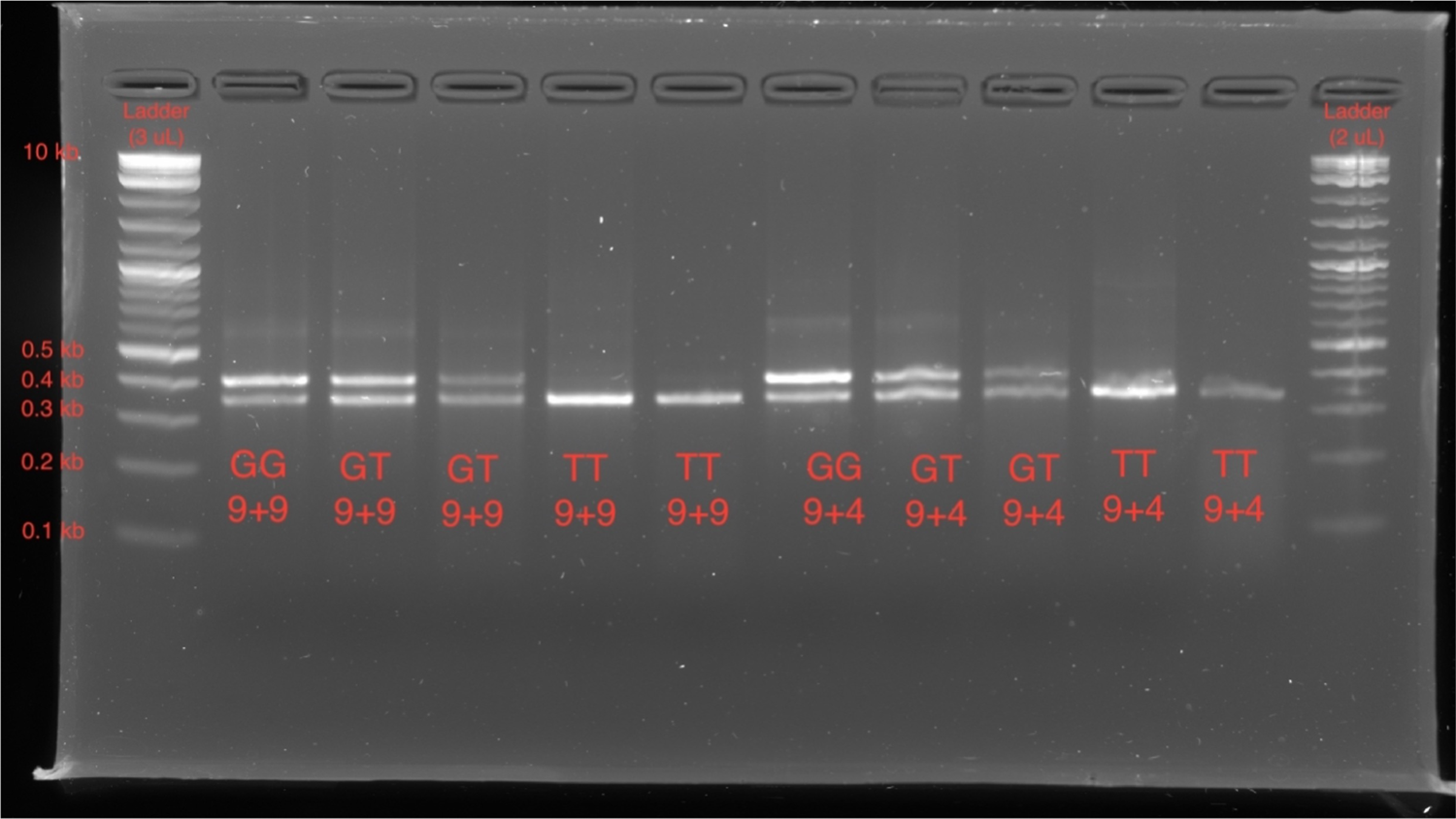
Reverse transcription PCR of RNA from lymphoblastoid cell lines of various genotypes (GG, GT, or TT). RNA was isolated from LCLs of various genotypes (GG, GT, or TT) and converted to cDNA followed by PCR amplification with primer pairs 9F and 9R (9+9) as well as 9F and 4R (9+4) (**Supplementary Table 7**). All PCR products were run on 1% agarose gel.

**Supplementary Figure 11:**
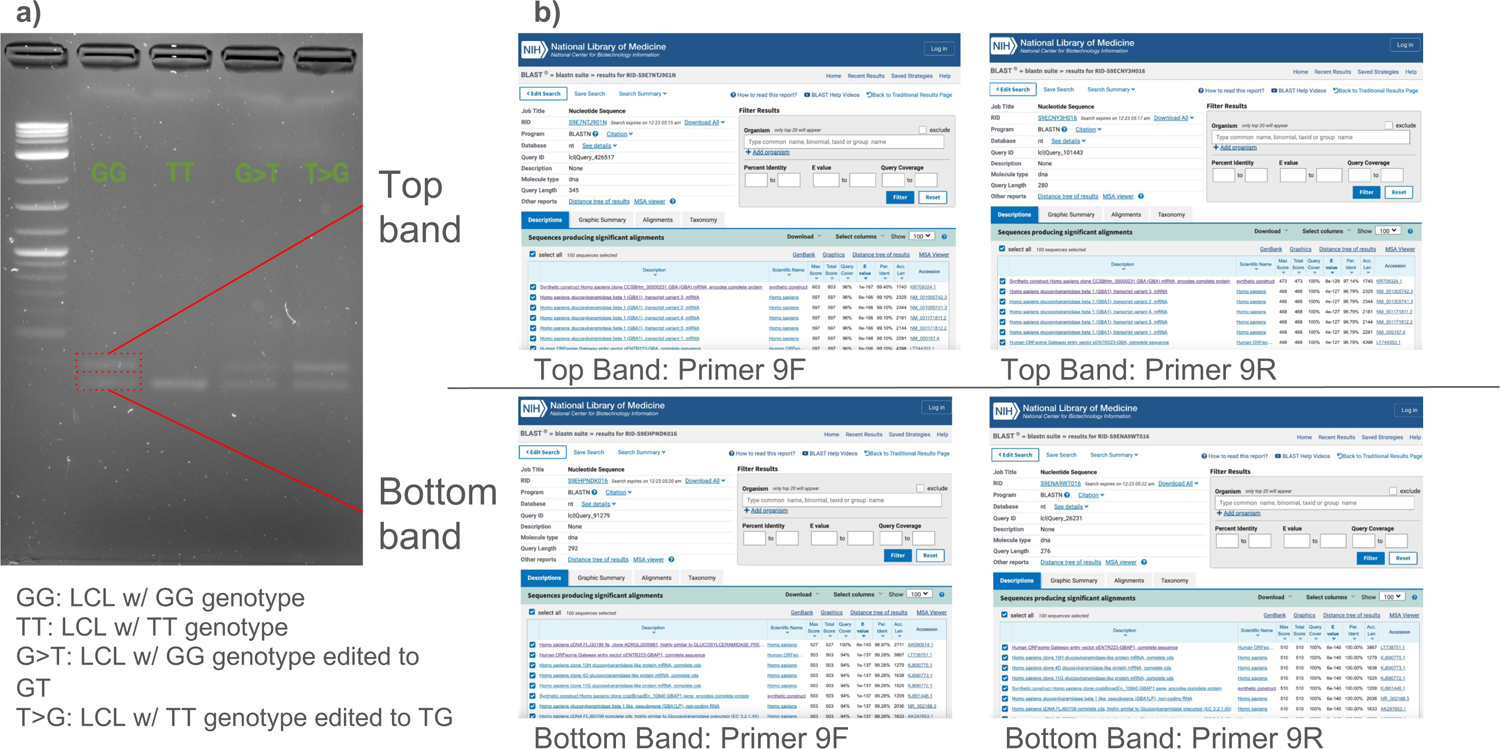
Reverse transcription PCR products used for Sanger sequencing. a) 1% agarose gel containing PCR products that were excised and sent for Sanger sequencing. b) BLAST results of sequences obtained from Sanger sequencing resulted in the top band to be *GBA1* transcript and the bottom band to be *GBAP1* showing the 55bp deletion difference between *GBA1* and *GBAP1* (See full BLAST results in **Supplementary Table 12**).

**Supplementary Figure 12:**
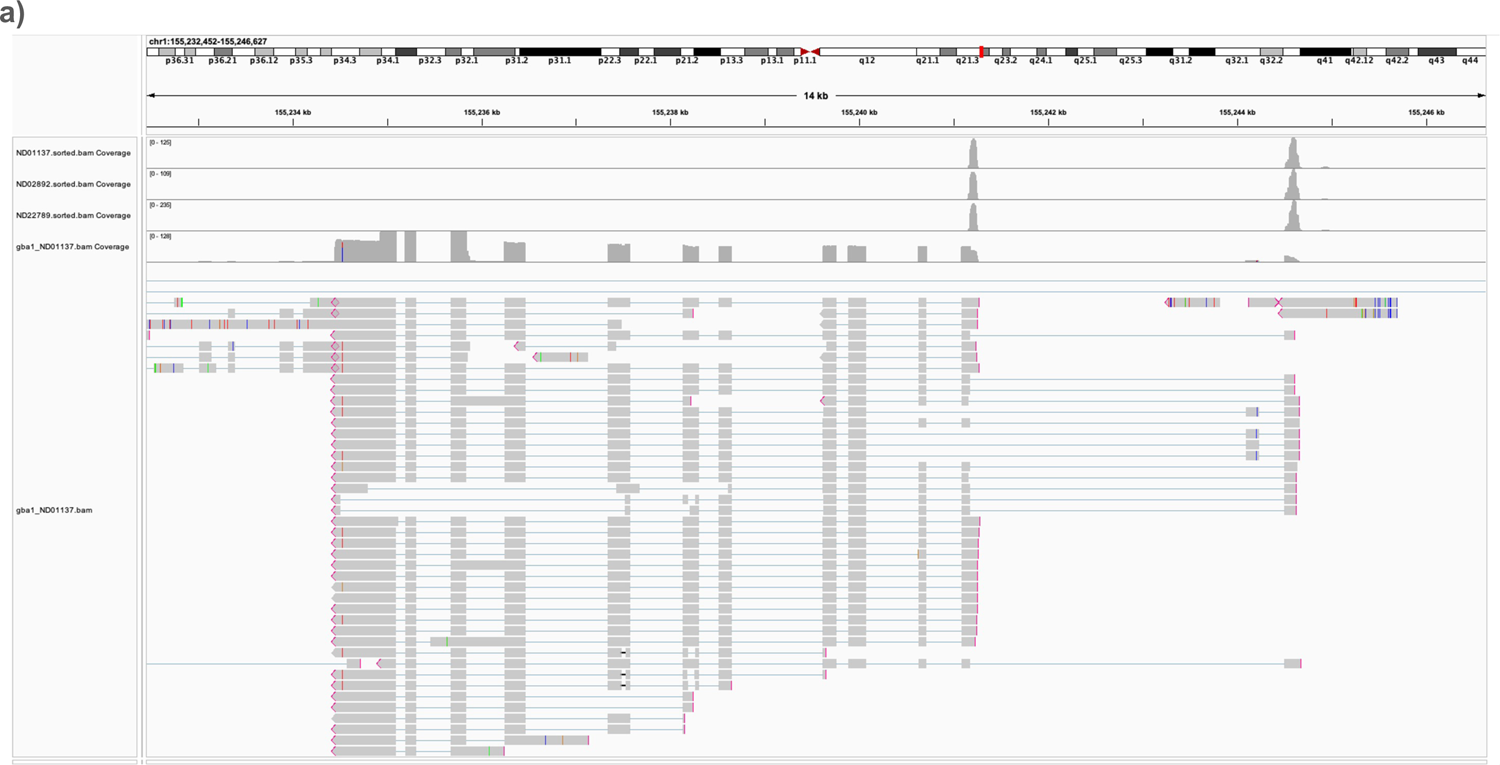
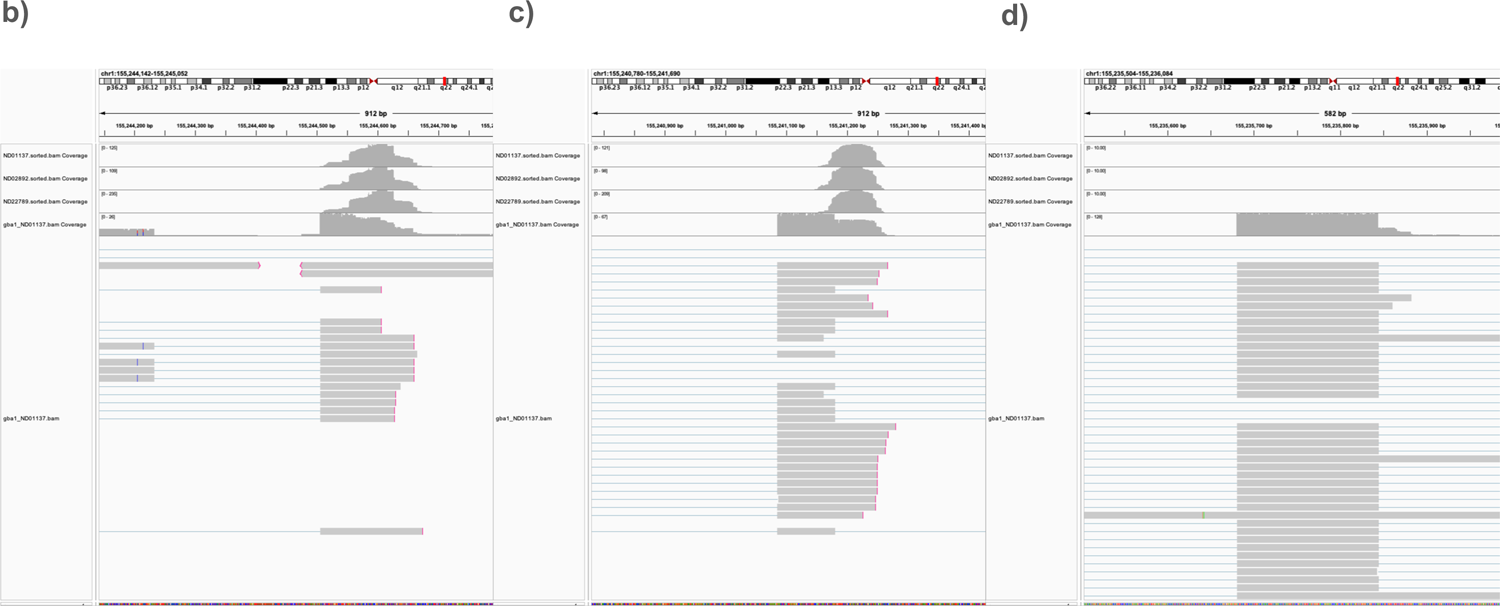
CAGEseq sequencing rs3115534-GG, GT, TT to assess transcription start sites across genotypes. a) Full view of the *GBA1* gene resulted in the identification of two transcription start sites at exons 1 and 2. No sequence reads were detected at the exon 9 region. b) zoomed in IGV screenshot of main transcription start site. c) zoomed in IGV screenshot of secondary transcription start site. d) zoomed in IGV screenshot of intron 8 region.

**Supplementary Figure 13:**
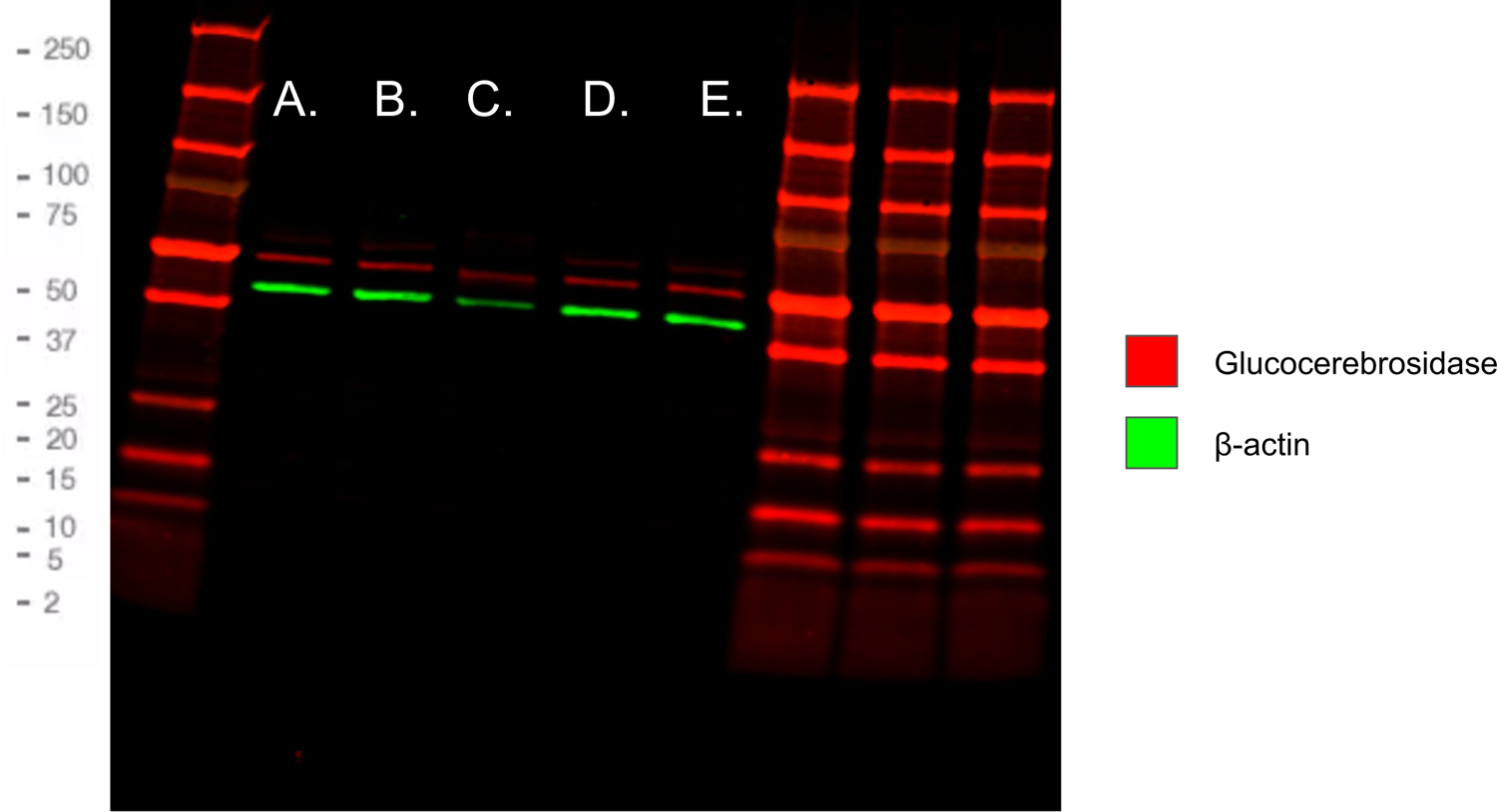
Western blot of various lymphoblastoid cell lines rs3115534 genotypes to assess protein coding ability. 30 ug of LCL protein lysate was loaded into each well. **A.** *GBA1* N370S Gaucher Disease Type 1 **B.** rs3115534-GG (replicate 1) **C.** rs3115534-GG (replicate 2) **D.** rs3115534-GT **E.** rs3115534-TT. Glucocerebrosidase (57.9 kD) is represented in red while β-actin (42 kD) is in green.

**Supplementary Figure 14:**
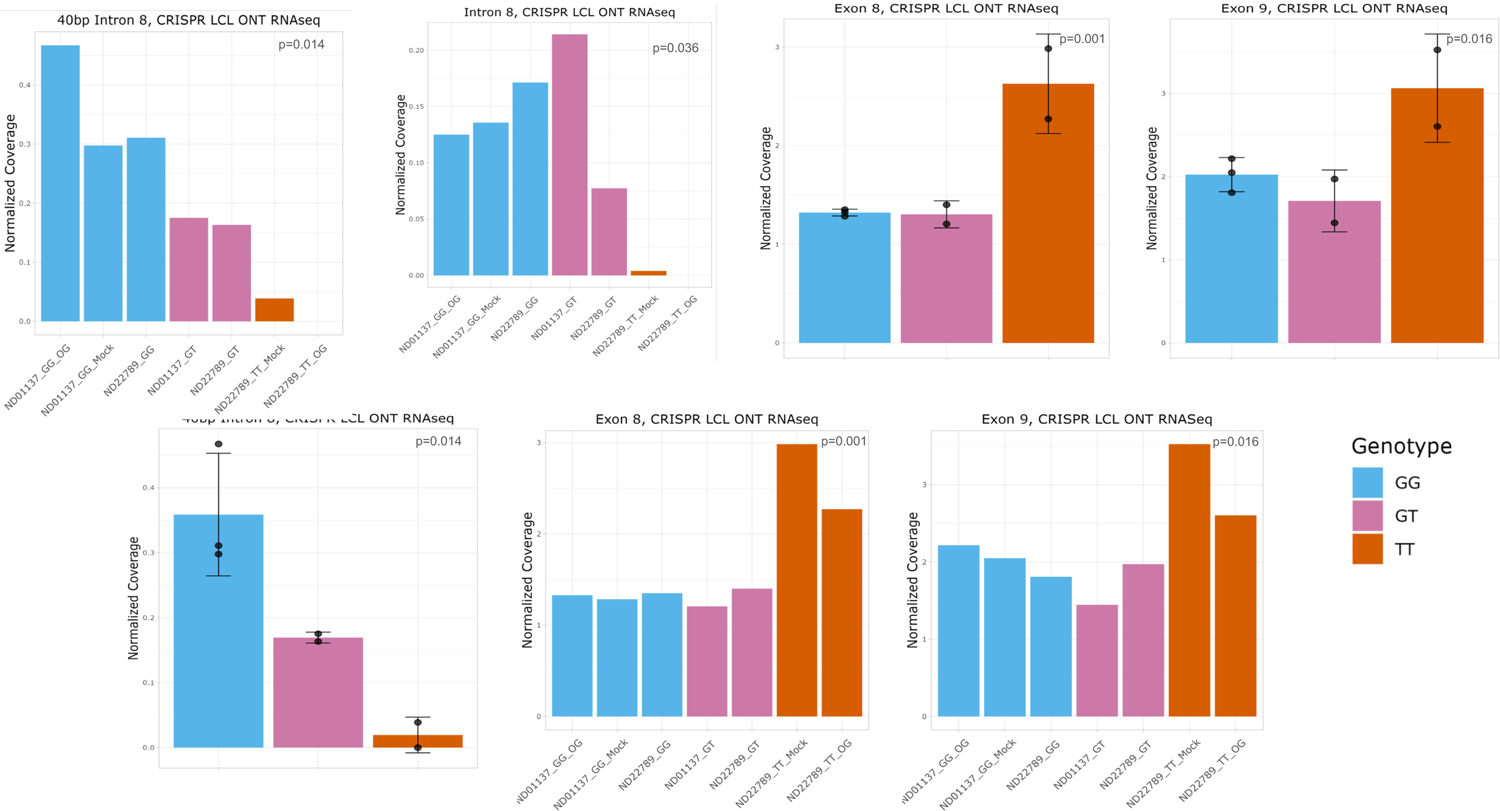
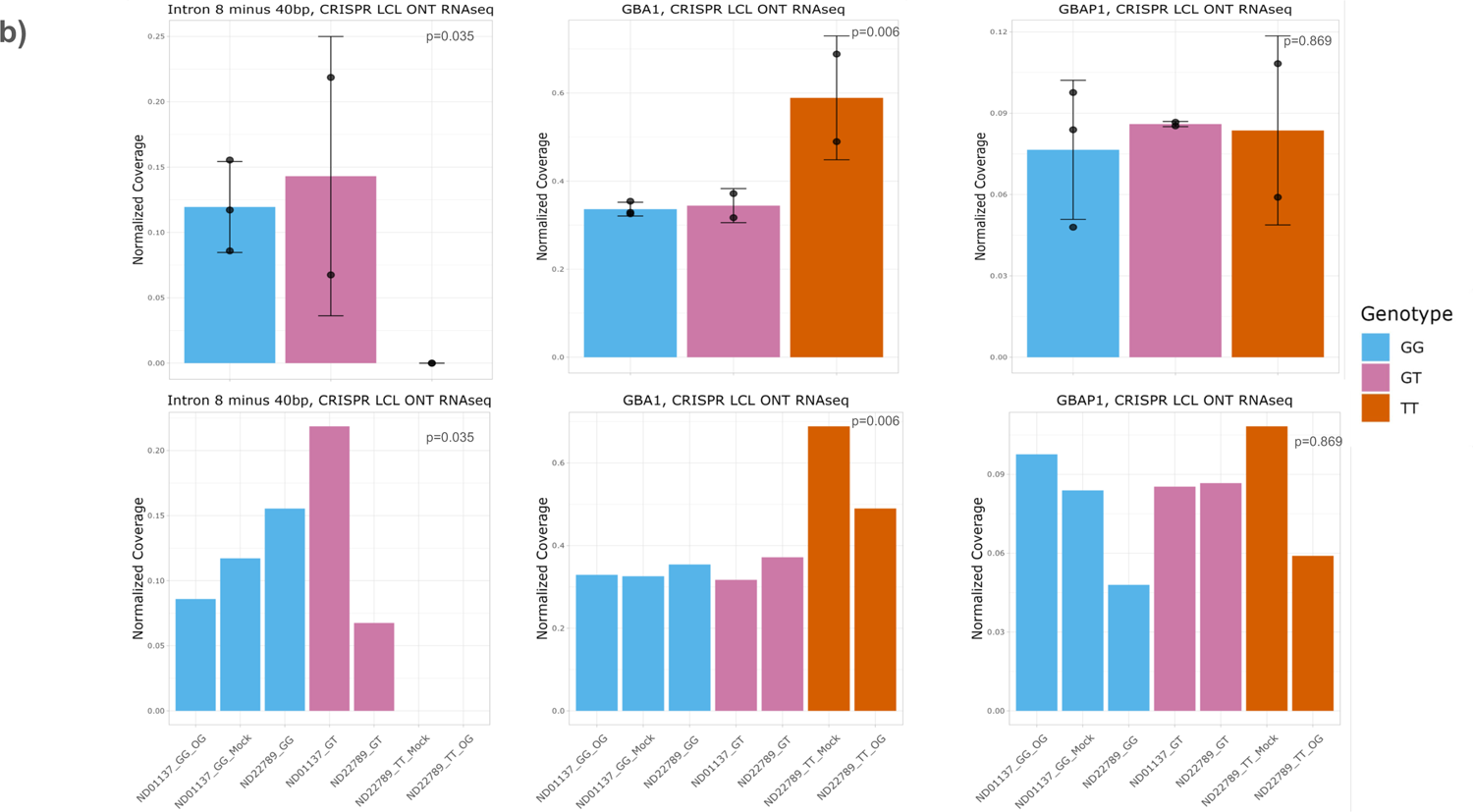
Additional coverage plots for CRISPR-edited lymphoblastoid cell lines sequenced with Oxford Nanopore Technologies long read RNA sequencing. a) Genotype collapsed quantification of intron 8 expression matching Figure 1 per sample plots plots. b) Additional coverage plots from CRISPR-edited LCLs for intron 8 minus 40bp transcript region, *GBA1*, and *GBAP1*. Plot are shown both per sample and collapse by genotype. Coverage for all panels normalized by dividing mean depth by total number of mapped reads per million as detailed in methods. Error bars represent standard deviation for all panels.

**Supplementary Figure 15:**
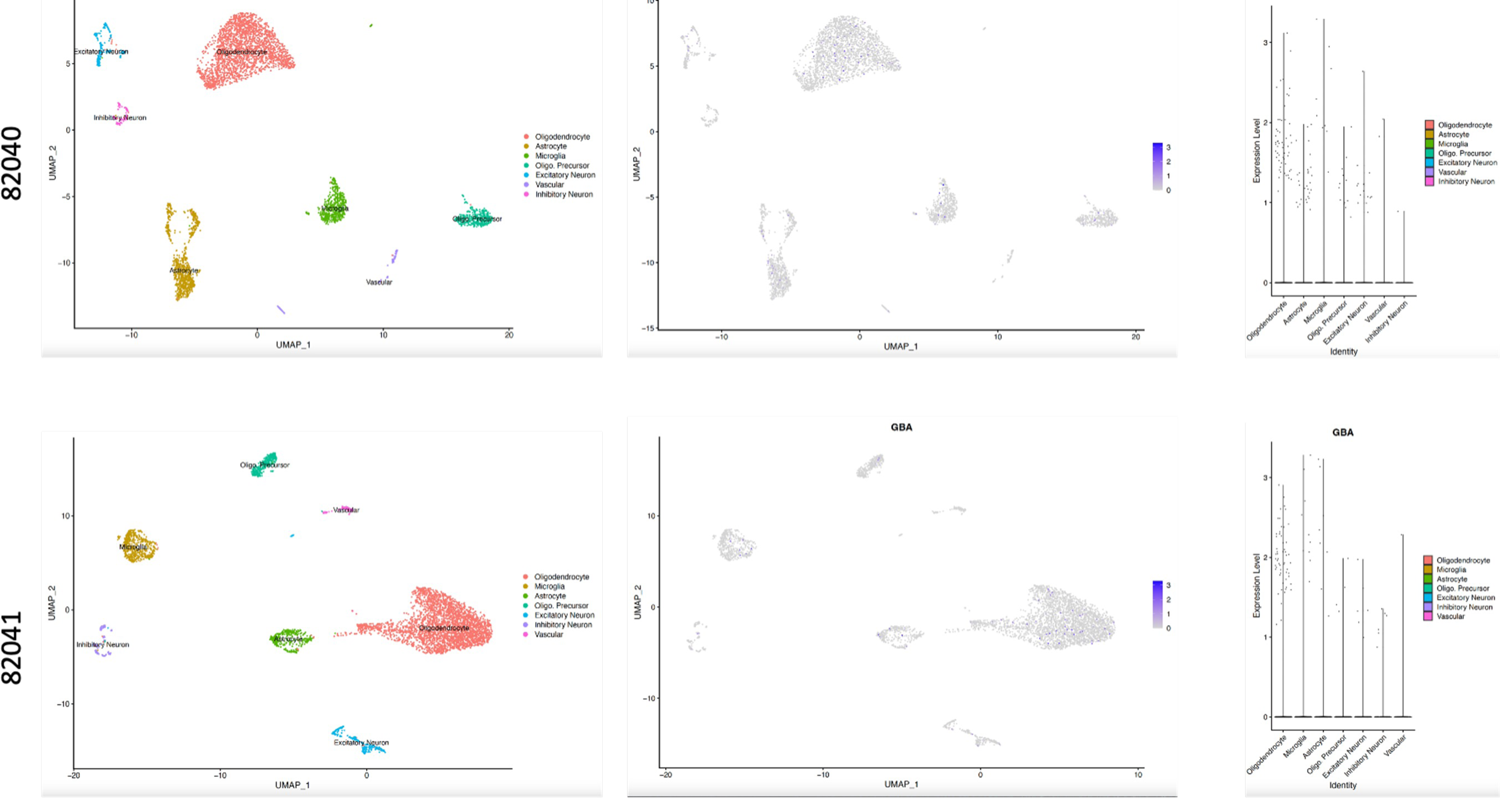
Low *GBA1* gene expression across major brain cell types using single nuclei RNA sequencing. UMAP of single nuclei RNA sequencing of two frontal cortex brain tissue samples (1x rs3115534-GG and 1x rs3115534-TT) from 10X Illumina data shows clear clustering of 7 major brain cell types. *GBA1* expression differences across cell types can also be picked up, however *GBA1* expression is generally low.

**Supplementary Figure 16:**
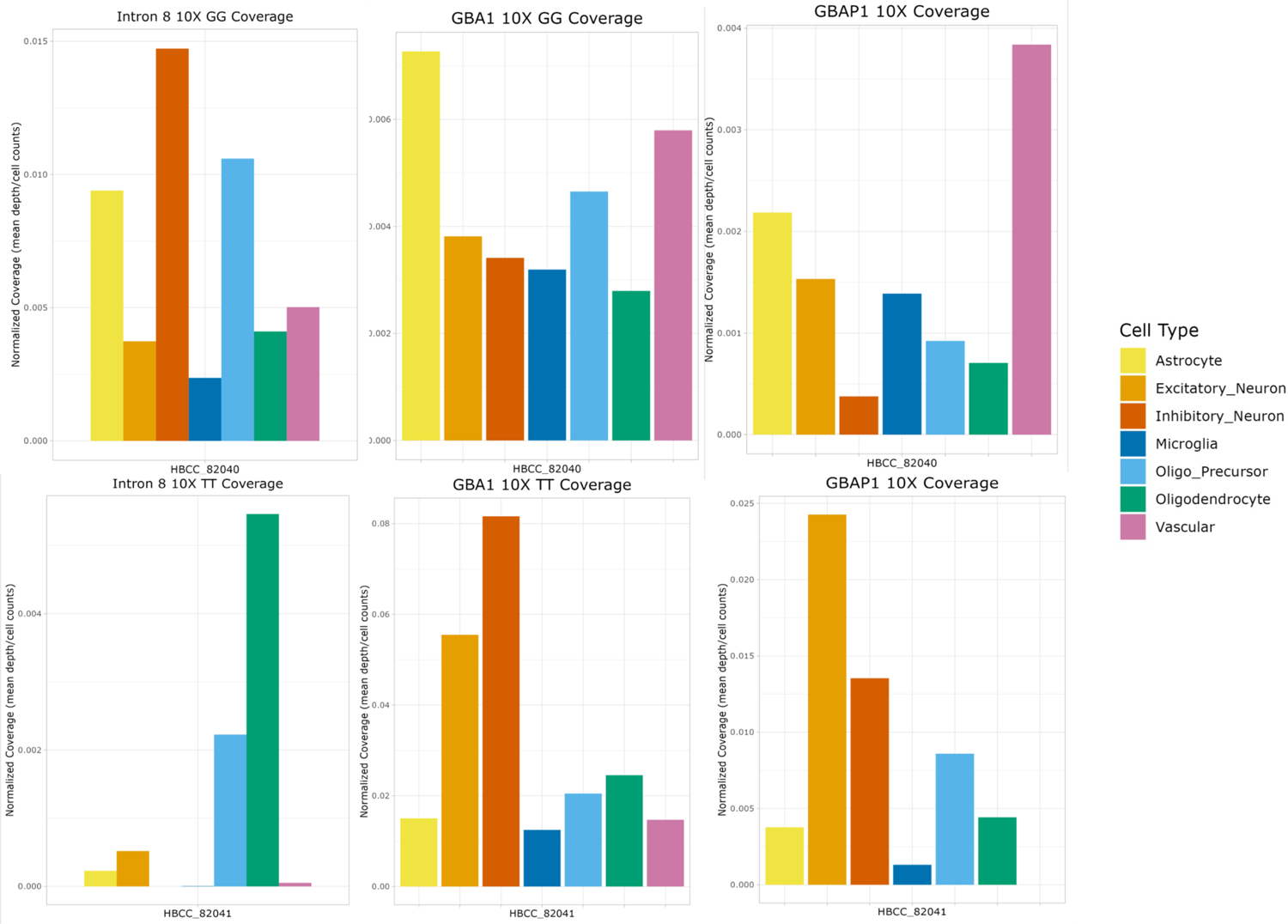
Enriching of *GBA1* in single nuclei RNA sequencing shows *GBA1* expression across major brain cell types. Enrichments of *GBA1* transcripts of two frontal cortex brain tissue samples (1x rs3115534-GG and 1x rs3115534-TT) using probes resulted in a clear increase of transcripts to be identified. Comparisons between rs3115534-GG and rs3115534-TT carriers showed no major difference between cell types between *GBA1* and *GBAP1*, however major differences were observed between GG and rs3115534-GG and rs3115534-TT in reads covering intron 8.

**Supplementary Figure 17:**
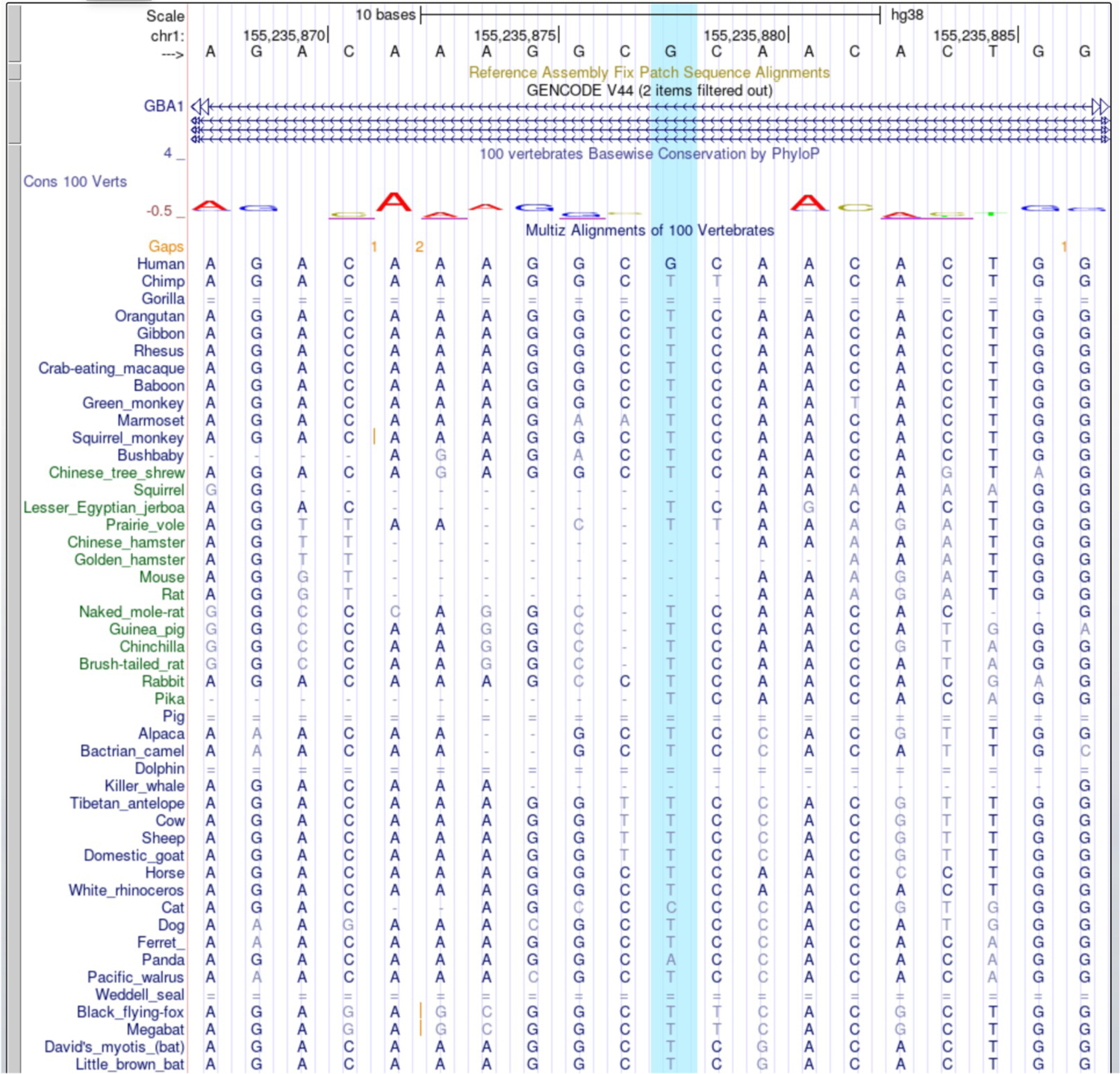
Assessment of conservation across species for rs3115534. The UCSC Genome Browser was accessed to investigate the conservation of this allele across vertebrates. Very high conservation was observed for the T allele in this part of the genome and humans are the only ones reported to have a G allele here as reference allele.

**Supplementary Figure 18:**
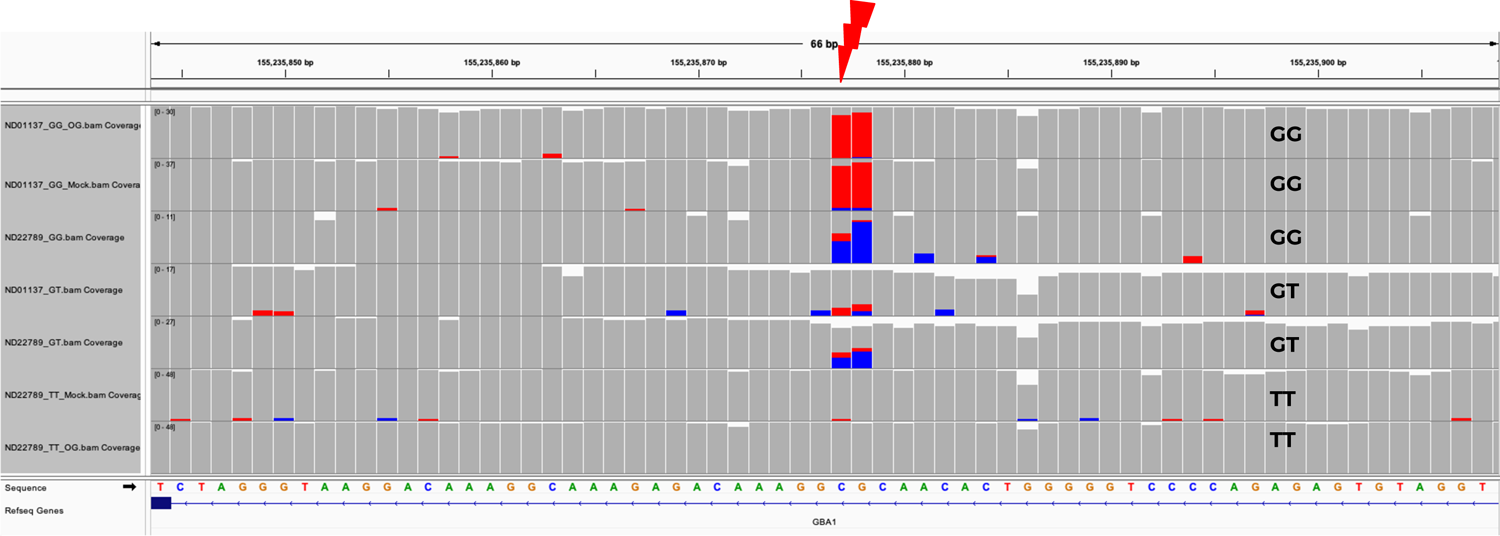
Methylation differences in rs3115534 across CRISPR-edited lymphoblastoid cell lines. Methylation information obtained from Oxford Nanopore Technologies DNA sequencing. When the rs3115534-G risk allele is present, the locus turns into a methylation site. When rs3115534-T non-risk allele is present, the methylation site is lost.

**Supplementary Figure 19:**
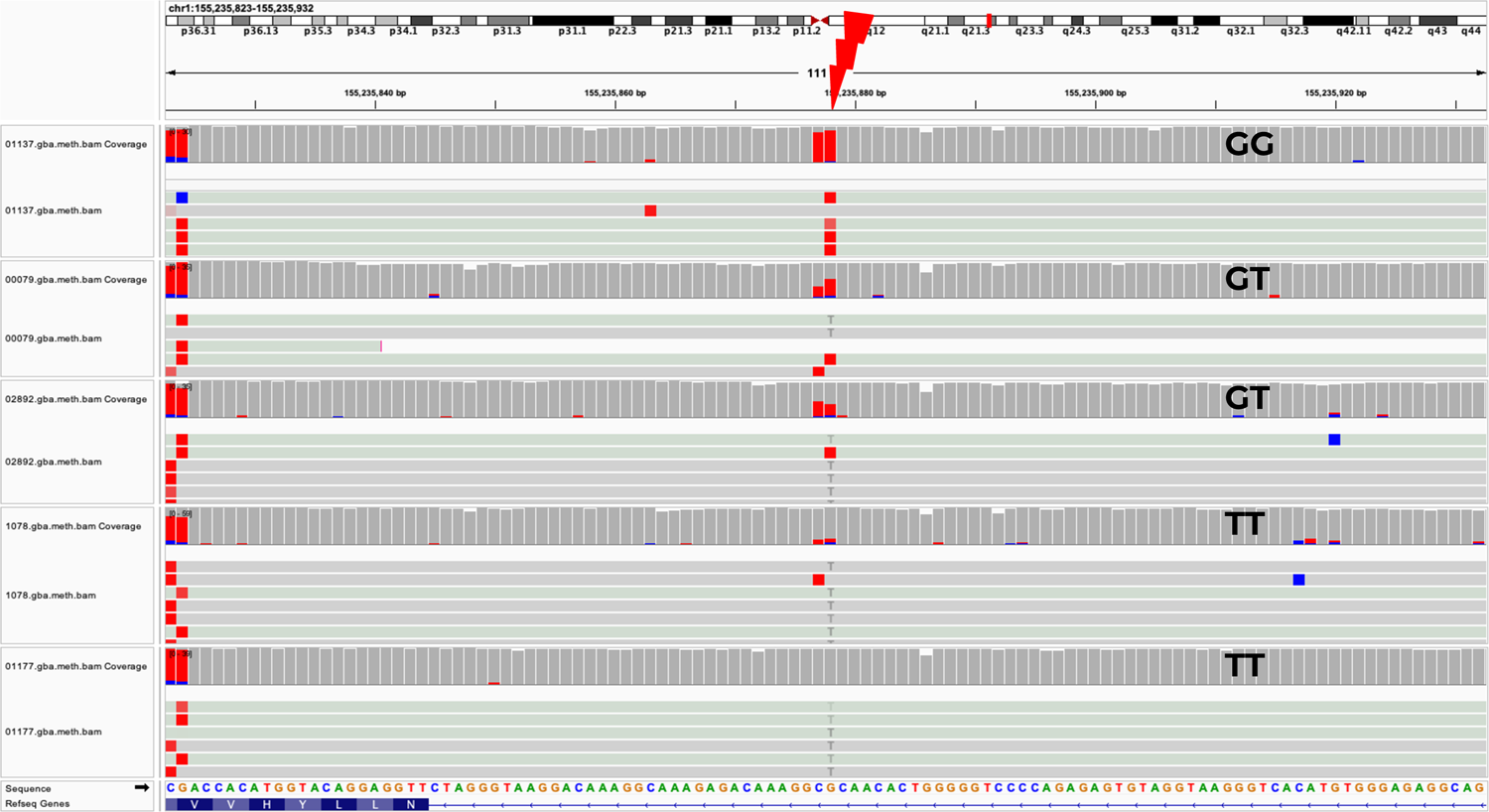
Methylation differences in rs3115534 across lymphoblastoid cell lines. Methylation information obtained from Oxford Nanopore Technologies DNA sequencing. When the rs3115534-G risk allele is present, the locus turns into a methylation site. When T-non-risk allele is present, the methylation site is lost.

**Supplementary Figure 20:**
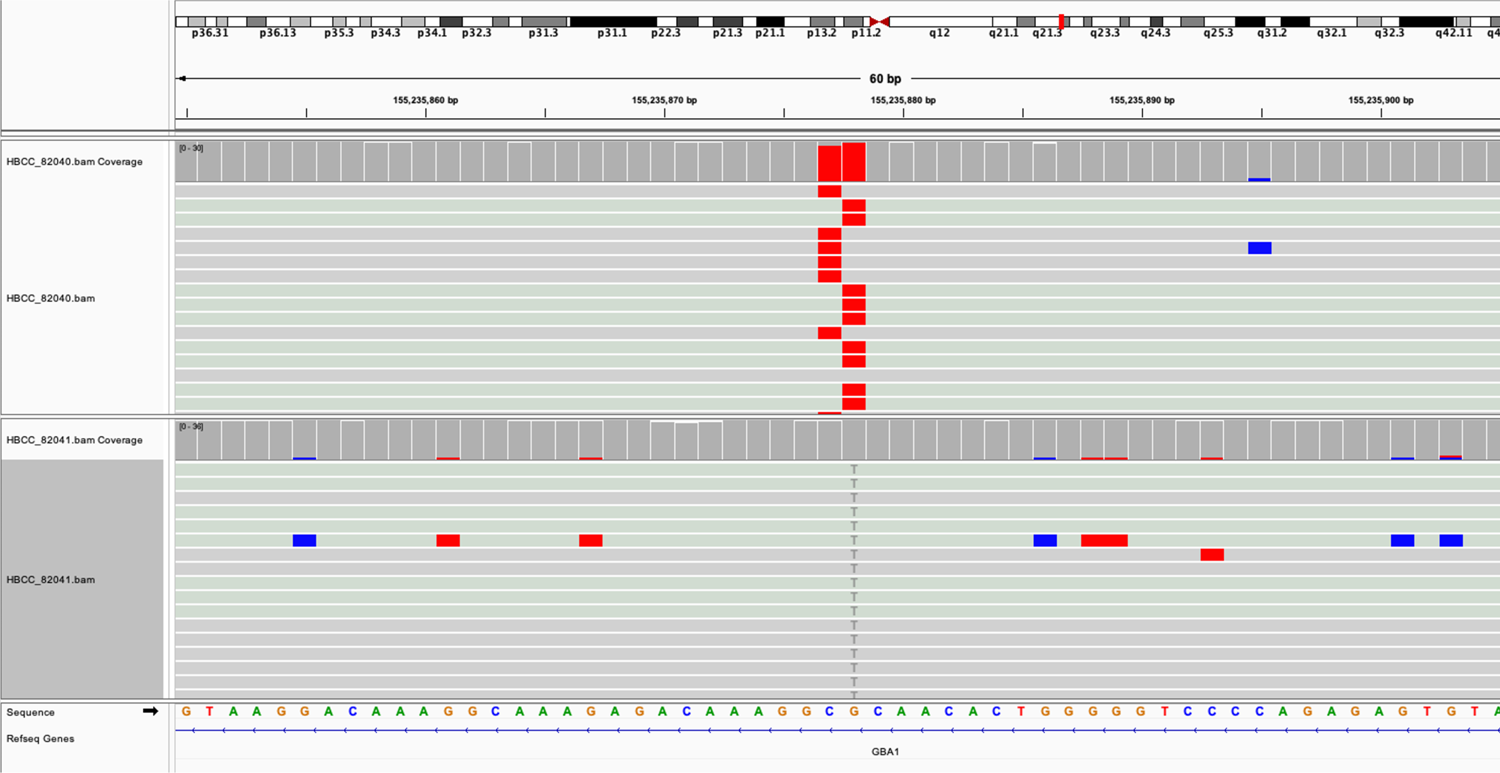
Methylation differences in rs3115534 across frontal cortex samples. Methylation information obtained from Oxford Nanopore Technologies DNA sequencing. HBCC_82040 is rs3115534-GG (top track), while HBCC_82041 (bottom track) is rs3115534-TT. When the rs3115534-G risk allele is present, the locus turns into a methylation site. When T-non-risk allele is present, the methylation site is lost.

**Supplementary Figure 21:**
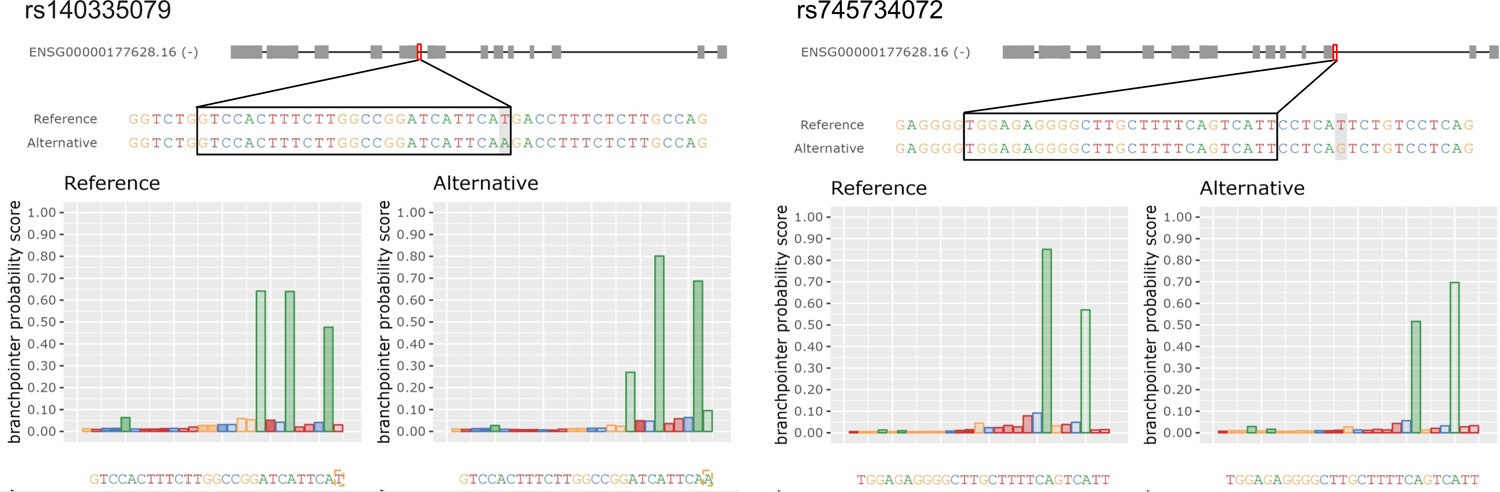
Branchpoint investigation of rs140335079 and rs745734072. rs140335079 is reported by AGAIN to be in a branchpoint sequence. The A is the reference allele and T is the alternative allele. No drastic differences were observed in the Branchpointer probability scores. rs745734072 is reported by AGAIN to be in a branchpoint sequence. The A is the reference allele and C is the alternative allele. No drastic differences were observed in the Branchpointer probability scores.

**Supplementary Figure 22:**
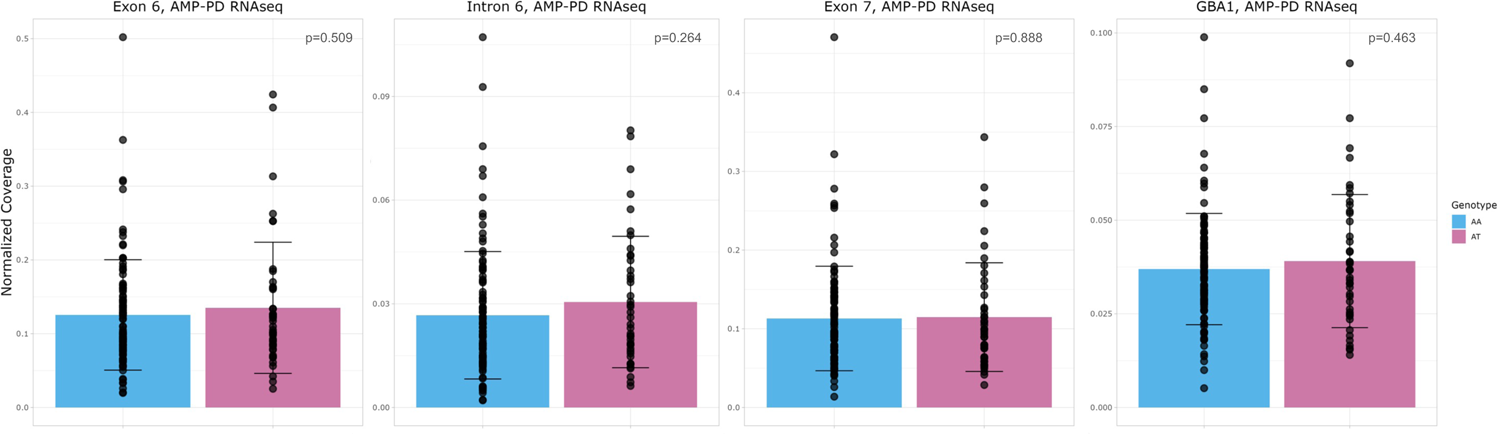
No expression differences identified due to rs140335079. rs140335079 is reported by AGAIN to affect a branchpoint sequence, however no drastic differences were observed in the Branchpointer probability scores. To confirm that the was no similar branchpoint disruption mechanism like in rs3115534, expression around rs140335079 was plotted using AMP-PD blood-based RNAseq data (n=146) and no important intron retention events were observed based on genotype.

## Supplementary Tables

**Supplementary Table 1: Biosamples used for assessment of effects of *GBA1* rs3115534.** Lymphoblastoid cell lines (LCLs) from Coriell Institute for Medical Research and frontal cortex tissue samples from the Human Brain Collection Core of GG-GT- and TT-rs3115534 genotypes were used. Sample type (LCLs or front cortex), origin, genotype, RNA Integrity numbers (RIN), and downstream analyses are provided for each sample.

**Supplementary Table 2: Probes used to capture *GBA1* transcripts.** Custom designed *GBA1* xGen Lockdown Probes (IDT) were used to specifically select and subsequently amplify any *GBA1* cDNA prior to sequencing.

**Supplementary Table 3: 10x ONT capture methods.** The ONT capture protocol consisted of an initial cDNA amplification, followed by hybridization with custom probes to capture *GBA1* cDNA. The captured cDNA was then amplified before proceeding to library preparation. During library preparation captured and amplified cDNA underwent biotin tagging and pull-down PCR before sequencing.

**Supplementary Table 4: Bed file for coverage plots of *GBA1*.** Ensembl coordinates were used to assess coverage in various *GBA1* transcript regions including the ∼40 bp novel transcript region observed in intron 8, both exons 8 and 9, intron 8; intron 8 excluding the ∼40 bp novel transcript, and lastly both *GBA1* and *GBAP1*.

**Supplementary Table 5: AMP-PD samples used for coverage information.** Data was accessed through the AMP-PD platform for WGS and RNAseq. Individual sample identifiers, case-control status, sex, predicted ancestry, rs3115534 variant status and cohort or origin are provided.

**Supplementary Table 6: 1000 Genomes samples used for coverage information.** Data was accessed through The International Genome Sample Resource platform for WGS and RNAseq. Individual sample identifiers, sex, ancestry and specific population, as well as rs3115534 variant status are provided.

**Supplementary Table 7:** HBCC Illumina RNAseq samples used for coverage information. Data was accessed through the Human Brain Collection Core for Illumina RNAseq data. Individual sample identifiers, disease status, and rs3115534 variant status are provided.

**Supplementary Table 8: Reverse transcription PCR Primers and Conditions.** Primers 9F, 9R and 4R were selected after initial optimization–respective sequences are provided. In an effort to increase specificity a touchdown PCR method was used as detailed in the PCR conditions.

**Supplementary Table 9: Sanger sequencing of excised PCR bands.** Sanger sequencing results were visually inspected and select regions were searched in the publically available BLAST database. The best matched BLAST result is listed along with both complete and selected sequences and a total base pair count for each sequence that is provided.

**Supplementary Table 10: CRISPR edited LCLs for rs3115534.** All original LCLs were obtained from Coriell. CRISPR editing was performed by Synthego. ND01137 was originally rs3115534-GG while ND22789 was originally rs3115534-TT. Both lines were mock transfected, partially edited to yield heterozygote pools, and completely edited to homozygotes for the inverse allele. Note that complete editing of ND01137 to rs3115534-TT was not successful and genotyping revealed a heterozygous cell pool. Downstream analyses performed are listed by line.

**Supplementary Table 11: Overview of statistical tests performed for coverage.** A variety of statistical tests were performed to compare coverage for various sequence regions from multiple datasets including, 1000Genomes LCL, AMP-PD Blood, HBCC Frontal Cortex ONT, HBCC Frontal Cortex Illumina, Coriell Illumina LCL, Coriell ONT LCL, CRISPR Coriell ONT LCL, AMP-PD Blood.

**Supplementary Table 12: Transcript quantifications using Stringtie2.** Normalized transcript counts for original LCLs, CRISPR edited LCLs, and HBCC brain samples are shown. COV-coverage; FPKM-fragments per kilobase of transcript per million mapped reads; TPM-transcripts per million; RATIO-novel transcript TPM divided by main transcript TPM; PRIMARY ALIGNED READS: WHOLE-all primary aligned read counts whole transcriptome; GBA-count of primary reads aligned to *GBA1*.

**Supplementary Table 13:** Mass spectrometry analysis of excised 4-20% agarose gel region predicted to contain truncated *GBA1* protein. rs3115534-GG (replicate 1) and (replicate 2) represent cell lysates from the same stock vial of ND01137 LCLs electrophoresed in parallel on the same gel and excised at the same time. These correspond to lanes B and C of Supplementary Figure 11. The remaining samples rs3115534-GT and rs3115534-TT correspond to lanes D and E, respectively. No known *GBA1* motifs were detected. # PSM-peptide-spectrum match number.

**Supplementary Table 14: Reads mapping to *GBA1* after 10x capture enrichment.** All primary aligned reads mapping to *GBA1* or *GBAP1* after performing the 10x capture enrichment for *GBA1* are shown. For downstream analysis of cell type specificity, the number of primary aligned reads with 10x barcodes was also quantified.

**Supplementary Table 15: *GBA1* variants affecting branchpoint sequences according to AGAIN algorithm.** List of three variants of interest in *GBA1* obtained from gnomAD that are predicted to impact a branchpoint sequence. See Figure 4 and Supplementary Figure 21 for a depiction of the scale of branchpoint sequence impact of these variants. CHR-chromosome; POS-base pair; ID-Chr:Pos:Ref:Alt; ID_RS-available rs ID; REF-reference allele.

## Data Availability

All generated LCL Coriell ONT DNAseq, CAGEseq and RNAseq data (ILM and ONT) is available at https://www.amp-pd.org/ via GP2 tier 2 access. AMP-PD ILM blood based RNAseq is available at https://www.amp-pd.org/ after signing the data use agreement. 1000 Genomes project data is publicly available at https://www.internationalgenome.org/. All generated brain tissue bulk RNAseq data (ONT) is currently being submitted to the NIMH data sharing platform at https://nda.nih.gov/. Summary statistics for cis-eQTLs and a catalog of ancestry-specific eQTLs from Kachuri et al.^12^ were obtained from https://doi.org/10.5281/zenodo.7735723.

## Acknowledgements

We would like to thank all of the participants who donated their time and biological samples to be a part of this study. This work utilized the computational resources of the NIH HPC Biowulf cluster (http://hpc.nih.gov). We also want to thank the Coriell cell repository and 1000 Genomes without their valuable resources including the genetic diversity presence this work would not be possible. The tissue used in this research was obtained from the Human Brain Collection Core, Intramural Research Program, NIMH (http://www.nimh.nih.gov/hbcc). Protein QTL data was generated using NIDDK grant number: R01DK108803. This project was supported by the Global Parkinson’s Genetics Program (GP2). GP2 is funded by the Aligning Science Across Parkinson’s (ASAP) initiative and implemented by The Michael J. Fox Foundation for Parkinson’s Research (https://gp2.org). For a complete list of GP2 members see https://gp2.org. This research has been conducted using the UK Biobank Resource under Application Number 33601. Data used in the preparation of this article were obtained from the Accelerating Medicine Partnership® (AMP®) Parkinson’s Disease (AMP PD) Knowledge Platform. For up-to-date information on the study, visit https://www.amp-pd.org. The AMP® PD program is a public-private partnership managed by the Foundation for the National Institutes of Health and funded by the National Institute of Neurological Disorders and Stroke (NINDS) in partnership with the Aligning Science Across Parkinson’s (ASAP) initiative; Celgene Corporation, a subsidiary of Bristol-Myers Squibb Company; GlaxoSmithKline plc (GSK); The Michael J. Fox Foundation for Parkinson’s Research; Pfizer Inc.; AbbVie Inc.; Sanofi US Services Inc.; and Verily Life Sciences. ACCELERATING MEDICINES PARTNERSHIP and AMP are registered service marks of the U.S. Department of Health and Human Services.

PPMI is sponsored by The Michael J. Fox Foundation for Parkinson’s Research and supported by a consortium of scientific partners: 4D Pharma, AbbVie Inc., AcureX Therapeutics, Allergan, Amathus Therapeutics, Aligning Science Across Parkinson’s (ASAP), Avid Radiopharmaceuticals, Bial Biotech, Biogen, BioLegend, BlueRock Therapeutics, Bristol Myers Squibb, Calico Life Sciences LLC, Celgene Corporation, DaCapo Brainscience, Denali Therapeutics, The Edmond J. Safra Foundation, Eli Lilly and Company, Gain Therapeutics, GE Healthcare, GlaxoSmithKline, Golub Capital, Handl Therapeutics, Insitro, Janssen Pharmaceuticals, Lundbeck, Merck & Co., Inc., Meso Scale Diagnostics, LLC, Neurocrine Biosciences, Pfizer Inc., Piramal Imaging, Prevail Therapeutics, F. Hoffmann-La Roche Ltd and its affiliated company Genentech Inc., Sanofi Genzyme, Servier, Takeda Pharmaceutical Company, Teva Neuroscience, Inc., UCB, Vanqua Bio, Verily Life Sciences, Voyager Therapeutics, Inc., and Yumanity Therapeutics, Inc. The PPMI investigators have not participated in reviewing the data analysis or content of the manuscript. For up-to-date information on the study, visit www.ppmi-info.org.

The Parkinson’s Disease Biomarker Program (PDBP) consortium is supported by the National Institute of Neurological Disorders and Stroke (NINDS) at the National Institutes of Health. A full list of PDBP investigators can be found at https://pdbp.ninds.nih.gov/policy. The PDBP investigators have not participated in reviewing the data analysis or content of the manuscript.

Special thanks to Synthego for the CRISPR editing and thanks to Psomagen for sequencing RNAseq Illumina libraries and Sanger sequencing. We would like to thank Boma Fubara and Yan Li at the NINDS Proteomics Core Facility for Protein Identification Analysis for efficiently performing the mass spectrometry analysis. We would also like to thank Hannah Macpherson from the University of College of London for guidance during the *GBA1* single-nuclei capture protocol.

## Reagent Availability

Unedited Coriell LCL lines are available at https://www.coriell.org/. CRISPR edited Coriell LCL lines are available upon request and establishment of an MTA with Coriell and NIH/CARD abiding by the Coriell NINDS Human Genetics Repository Material Transfer Agreement For Biospecimens.

## Author contributions

Conceptualization: PAJ, PAWC, ChB, MW, NUO, CoB

Experiments were performed by: PAJ, PAWC, DMR, MR, KJB, LM, SB, CH, ZS, JS, YAQ, CE

Data was provided by: ALS, HLL, LK, ChB, PB, SF

Data was analyzed by: PAJ, PAWC, DR, EKG, MR, MBM, OOO, KJB, LM, KD, SB, CH, ZS, ALS, JS, MZ, HRM, CBP, HLL, LS, YAQ, MAN, SBC, JH, HH, CE, EGB, LK, ABS, SF, PB, XR, MR, ChB, MW, NUO, CoB

Writing (original draft): PAJ, PAWC,CoB

Writing (review and editing): PAJ, PAWC, DR, EKG, MR, MBM, OOO, KJB, LM, KD, SB, CH, ZS, ALS, JS, MZ, HRM, CBP, HLL, LS, YAQ, MAN, SBC, JH, HH, CE, EGB, LK, ABS, SF, PB, XR, MR, ChB, MW, NUO, CoB

## Code Availability

All scripts and code for this project can be found at: https://github.com/GP2code/GBA1-rs3115534-branchpoint

## Funding agencies

This work was supported in part by the Intramural Research Program of the National Institutes of Health including: the Center for Alzheimer’s and Related Dementias, within the Intramural Research Program of the National Institute on Aging and the National Institute of Neurological Disorders and Stroke (1ZIAAG000538-03, ZIAAG000542-01 and 1ZIAAG000543-01). GP2 is funded by the Aligning Science Across Parkinson’s (ASAP) initiative and implemented by The Michael J. Fox Foundation for Parkinson’s Research (https://gp2.org). This research was funded in part by Aligning Science Across Parkinson’s MJFF-024547 through the Michael J. Fox Foundation for Parkinson’s Research (MJFF).

The generation of molecular data for the TOPMed program was supported by the National Heart, Lung, and Blood Institute (NHLBI). RNA-seq for the NHLBI TOPMed Genes-Environments and Admixture in Latino Asthmatics Study (GALA II; phs000920) and Study of African Americans, Asthma, Genes, and Environments (SAGE; phs000921) was performed at the Broad Institute Genomics Platform (HHSN268201600034I). WGS for the same studies was performed at the NYGC (3R01HL117004-02S3) and NWGC (HHSN268201600032I). Core support including centralized genomic read mapping and genotype calling, along with variant quality metrics and filtering, was provided by the TOPMed Informatics Research Center (3R01HL-117626-02S1; contract HHSN268201800002I). Core support including phenotype harmonization, data management, sample identity quality control and general program coordination was provided by the TOPMed Data Coordinating Center (R01HL-120393 and U01HL-120393; contract HHSN268201800001I). WGS as part of GALA II was performed by the NYGC under a grant from the Centers for Common Disease Genomics of the GSP (UM1 HG008901). The GSP Coordinating Center (U24 HG008956) contributed to cross-program scientific initiatives and provided logistical and general study coordination. The GSP is funded by the National Human Genome Research Institute, NHLBI and National Eye Institute. This work and E.G.B. were supported in part by the Sandler Family Foundation, American Asthma Foundation, Robert Wood Johnson Foundation Amos Medical Faculty Development Program, Harry Wm. and Diana V. Hind Distinguished Professor in Pharmaceutical Sciences II, NHLBI (R01HL117004, R01HL135156, X01HL134589 and U01HL138626), National Institute of Environmental Health Sciences (R01ES015794), National Institute on Minority Health and Health Disparities (R56MD013312 and P60MD006902), Tobacco-Related Disease Research Program (24RT-0025 and 27IR-0030), and National Human Genome Research Institute (U01HG009080). This research was funded in part by Aligning Science Across Parkinsons [ASAP 000478].

## Ethics Statement

The specimens and data involved in this study were not collected specifically for this study and no one included in the study team had, nor currently has, access to the subject identifiers connected to the biospecimens or data, therefore, according to 45 CFR 46 per the Office for Human Research Protections under the Department of Health and Human Services, this study is not considered human subjects research and the need for IRB approval for this study has been waived. For additional clarity, we have provided details regarding consent originally obtained by the publicly available sources we utilized.

An informed consent form was completed by all participants and/or their legal guardians for any and all individuals who donated samples that were included in this study. IRB approval was provided by each enrolling institution.

All cohorts recruited to the GP2 initiative undergo comprehensive review of the relevant consent forms provided in the Operations and Compliance working group, thus, ensuring that all contributing sites and studies abided by the proper ethics guidelines set out by their institutional review boards, and all participants provided informed consent for inclusion in both their initial cohorts and subsequent studies within local legal constraints. All GP2 data is hosted in collaboration with the Accelerating Medicines Partnership in Parkinson’s disease, and is accessible via online application (https://amp-pd.org/register-for-amp-pd). Data from 1000 Genomes is publicly available via an online application (https://www.internationalgenome.org/) and all participants were required to fill out informed consents prior to participation. Data accessed from 1000 genomes was done so in compliance with proper guidelines. Populations included in the 1000 genomes collection as part of the IGSR have Consent, Ethics Review and Sampling Process which meet the criteria established for the 1000 Genomes Project and the applicants confirm to the P3G-IPAC that these criteria have been sought prior to sample collection, as was done by the Samples and ELSI subgroup for 1000 Genomes Project sample sets, only then does P3G-IPAC provide approval for acceptance for deposit in IGSR. All data obtained from UK Biobank was accessed following an online application (https://www.ukbiobank.ac.uk/) and project approval in compliance with UK Biobank data use guidelines and approved study protocols. UK Biobank has approval from the North West Multi-centre Research Ethics Committee (MREC) as a Research Tissue Bank (RTB) approval. This approval means that researchers do not require separate ethical clearance and can operate under the RTB approval (there are certain exceptions to this which are set out in the Access Procedures, such as re-contact applications). This RTB approval was granted initially in 2011 and it is renewal on a 5-yearly cycle: hence UK Biobank successfully applied to renew it in 2016 and 2021. UK Biobank will in due course apply for renewal effective in 2026. These renewal applications and approvals are shown on the website. Samples from the Coriell Institute for Medical Research (https://www.coriell.org/1/NIGMS/How-to-Order) were accessed following approval of a comprehensive application and project proposal which was approved by the appropriate bodies following proper guidelines. The following cell lines/DNA samples were obtained from the NIGMS Human Genetic Cell Repository at the Coriell Institute for Medical Research: [ND01137, ND00079, ND02892, ND02895, ND05171, ND12179, ND15296, ND24020, ND02896, ND04724, ND01177, ND06940, ND11329, ND13434, ND22789, ND24062, ND25221, ND01078]. An in-depth application was completed for access to Human Brain Collection Core samples which were accessed following approval by the Human Brain Collection Core (https://www.nimh.nih.gov/research/research-conducted-at-nimh/research-areas/research-support-services/hbcc). The brain tissues are obtained under protocols approved by the CNS IRB (NCT00001260), with the permission of the next-of-kin (NOK) through the Offices of the Chief Medical Examiners (MEOs) in the District of Columbia, Northern Virginia and Central Virginia. Additional samples were obtained from the University of Maryland Brain and Tissue Bank (contracts NO1-HD-4-3368 and NO1-HD-4-3383) (http://www.medschool.umaryland.edu/btbank/ and the Stanley Medical Research Institute (http://www.stanleyresearch.org/brain-research/).

We confirm that all participant consent was obtained and the appropriate institutional documentation has been stored properly. We also affirm that any participant identifiers, including identifiers for donated biosamples, that were included in our study were not known to anyone (e.g., patients, hospital staff, or participants themselves) beyond the research group and thereby, cannot be used to identify any individual.

